# MIRA-Net: A Cross-Cohort Representation Learning Framework for Parkinson’s Disease Classification Using Acoustic and Beta-Band MEG Biomarkers

**DOI:** 10.64898/2026.07.03.26357258

**Authors:** N Akhila, Asif Ekbal, Dipanjan Roy

## Abstract

Accurate diagnosis of Parkinson’s disease (PD) remains challenging due to substantial inter-subject variability and the absence of widely accessible, objective multimodal biomarkers. Although speech and magnetoencephalography (MEG) biomarkers have individually demonstrated strong discriminative potential, their joint utilization is constrained by the absence of subject-level paired datasets—a fundamental gap that has prevented cross-modal validation at the individual level. We argue that this makes cross-cohort representation learning not merely a pragmatic workaround, but the most realistic and clinically transferable framework for multimodal PD assessment. In real-world deployment, acoustic screening and neuroimaging biomarkers are acquired through separate clinical pathways and must be integrated across heterogeneous patient populations. To address this, we propose MIRA-Net (Modality-Invariant Residual Adversarial Network). This cross-cohort representation learning framework integrates acoustic speech features from four established UCI datasets (n = 193) with beta-band MEG biomarkers from the NatMEG-PD dataset (n = 127) for PD classification. MIRA-Net employs RF-SHAP feature selection, gradient-reversal-based domain adaptation, and supervised contrastive alignment to learn participant-independent, modality-invariant embeddings. The framework is evaluated under Rest, Go, and Passive task conditions against Early Fusion, Vanilla DANN, and Supervised Contrastive Learning baselines. MIRA-Net achieves a peak accuracy of 86.23% (Go condition, Stacking classifier) with AUC values exceeding 0.88 under repeated cross-validation, alongside sensitivity of 89.4% and specificity of 83.1%. Friedman tests confirm statistically significant performance differences among fusion strategies (p < 0.003 across all conditions). These results demonstrate that cross-cohort representation learning can extract robust disease-discriminative signatures without synchronized multimodal recordings, offering a practical pathway toward AI-assisted PD assessment in resource-constrained clinical settings.

## 1. Introduction

Parkinson’s disease (PD) is a progressive neurodegenerative disorder characterized by the gradual loss of dopamine-producing neurons in the substantia nigra [1]–[3]. This degeneration leads to motor impairments, including tremor, rigidity, bradykinesia, and postural instability, as well as a range of non-motor symptoms that can significantly affect quality of life. Early and accurate diagnosis remains challenging because clinical assessment often relies on subjective neurological examinations and symptom-based observations, which may vary considerably across individuals. Consequently, there is growing interest in developing objective computational approaches capable of identifying disease-specific biomarkers from physiological and behavioural signals [4], [5].

Among the various non-invasive biomarkers, speech has emerged as a promising modality for Parkinson’s disease assessment [6]–[8]. Vocal impairments often appear during the early stages of the disease and can be acquired conveniently without specialized clinical equipment. Alterations in phonation, articulation, and speech rhythm reflect underlying neuromuscular dysfunction and have been widely exploited by machine learning and deep learning approaches for automated Parkinson’s disease detection [7]–[10]. However, speech-based biomarkers primarily capture the behavioural manifestations of the disease and may not fully reflect the underlying neural dysfunction responsible for motor impairment.

Magnetoencephalography (MEG) provides direct measurements of neuronal activity with excellent temporal resolution [11], [12] and offers valuable insights into the functional abnormalities associated with Parkinson’s disease. In particular, oscillatory activity within the beta frequency band (approximately 13–30 Hz) has been consistently associated with Parkinsonian motor dysfunction [13]–[16], with excessive beta-band synchronization linked to impaired movement initiation, bradykinesia, and rigidity. Therapeutic interventions such as dopaminergic medication and deep brain stimulation have also been shown to suppress pathological beta oscillations [14], [15], [17] and improve motor function. Consequently, beta-band MEG activity represents a physiologically meaningful biomarker for Parkinson’s disease and provides complementary neural information to acoustic biomarkers.

Although speech and MEG have each demonstrated considerable potential for Parkinson’s disease assessment, their joint utilization remains relatively underexplored [18]. Furthermore, practical deployment is challenged by the limited availability of subject-level multimodal datasets, variations in acquisition protocols across independent cohorts, and substantial inter-subject variability. Conventional approaches based on direct feature concatenation or standard domain adaptation techniques may therefore struggle to effectively exploit the complementary information contained in heterogeneous modalities while simultaneously addressing cross-cohort discrepancies. We note that the absence of subject-level paired recordings is not merely a data limitation to be overcome, but rather reflects the operational reality of clinical neurology: speech assessments and MEG recordings are acquired in distinct facilities, across separate appointments, and often in geographically distributed cohorts. This cross-cohort scenario is therefore the ecologically valid setting for multimodal PD biomarker research. Crucially, the shared disease label space across both modalities — PD versus healthy control — provides a principled semantic anchor for cross-modal alignment, enabling the learning of disease-discriminative representations that are invariant to modality origin and acquisition context. This motivates the design of MIRA-Net as a cross-cohort representation learning framework rather than a conventional subject-matched fusion approach.

Motivated by these challenges, we propose MIRA-Net, a cross-cohort representation learning framework that integrates acoustic and beta-band MEG biomarkers for Parkinson’s disease classification. First, RF-SHAP is employed to identify compact yet highly discriminative feature representations from both modalities. Subsequently, cross-modal correlation analysis is performed to quantify the degree of shared and complementary information captured by the selected acoustic and MEG features. Guided by these observations, MIRA-Net is designed to learn robust cross-cohort representations that preserve disease-discriminative characteristics while reducing modality-specific and participant-specific variability, thereby improving generalization across heterogeneous cohorts.

## 2. Proposed Methodology

In this section, we present the proposed MIRA-Net (Modality-Invariant Residual Adversarial Network) framework designed to address the challenges of cross-domain and cross-participant variability in Parkinson’s disease classification. The method integrates two complementary biomarkers: acoustic speech features and MEG derived neural features, to enhance diagnostic robustness across heterogeneous data sources. The overall framework is structured as follows.

### 2.1 Heterogeneous Datasets and Feature Spaces

The proposed framework was evaluated using heterogeneous acoustic and MEG datasets comprising 320 participants (193 acoustic and 127 MEG participants). The acoustic modality was constructed from four publicly available UCI Parkinson’s disease datasets, whereas the neural modality was obtained from the NatMEG-PD dataset. The NatMEG-PD dataset additionally provides MRI scans; however, only MEG recordings were utilized in this study. Participants with missing passive-movement recordings (n = 7) were excluded, resulting in a final cohort of 127 subjects. A summary of all datasets with their features is provided in **Table 1**.

**Table 1.** Overview of Acoustic and MEG Datasets and Feature Sets for Parkinson’s Disease Analysis.

| Modality | Dataset | Participants (PD/HC) | Instances | Feature Categories | Description |
| --- | --- | --- | --- | --- | --- |
| Acoustic | Max Little PD Dataset (2008) | 31 (23/8) | 195 | 22 acoustic features (Frequency, Jitter, Shimmer, Voice Quality) | Sustained vowel phonation recordings with clinically validated PD/HC labels [19]. |
|  | Multiple Sound Types Dataset (2013) | 40 (20/20) | 1040 | 29 acoustic features (Frequency, Jitter, Shimmer, Spectral, UPDRS) | Voice recordings with acoustic features and corresponding clinical scores [20]. |
|  | Replicated Acoustic Features Dataset (2019) | 80 (40/40) | 240 | 48 acoustic features (Frequency, Jitter, Shimmer, Perturbation, Spectral, Voice Quality) | Replicated acoustic measurements for PD classification [21]. |
|  | Telemonitoring Dataset (2009) | 42 (42/0) | 5875 | 22 acoustic features (Frequency, Jitter, Shimmer, UPDRS) | Longitudinal home-based speech recordings for PD monitoring [22]. |
| MEG | NatMEG-PD v1.1 (2023)-Resting state | 127 (62/65) | 7350 | 21 MEG features (Burst, Envelope, Spectral) | Resting-state (participants sat still with their eyes closed for approximately three minutes while spontaneous neural activity was captured) [23]. |
|  | NatMEG-PD v1.1 (2023)-Go task | 127 (62/65) | 7350 | 21 MEG features (Burst, Envelope, Spectral) | Go-task (participants responded to a visual colour-change cue by pressing a button as quickly as possible, across 150 trials, with short breaks between blocks) [23]. |
|  | NatMEG-PD v1.1 (2023)-Passive task | 127 (62/65) | 7350 | 21 MEG features (Burst, Envelope, Spectral) | Passive-task (a mechanical actuator moved the participant's index finger while they remained relaxed and refrained from voluntary movement across 150 trials) [23]. |

### 2.2 Feature Harmonization and Selection

Detailed feature descriptions are provided in the Supplementary Material. Due to variations in recording conditions and participant populations, each acoustic dataset was considered as a distinct cohort [24], [25]. The four acoustic datasets were harmonized through schema alignment and merged into a unified repository [24], [26]. After removing metadata attributes, the resulting acoustic dataset contained 7,350 recordings represented by 70 acoustic features. RF and SHAP were jointly employed to identify the most discriminative predictors [27]-[29], reducing the feature space to seven features (7,350 × 7). For the MEG modality, 21 beta-band features were extracted from 306 sensors [30], [31]. A graph-based channel selection strategy based on Pearson correlation and degree centrality identified representative sensors [32], [33], yielding a harmonized dataset of size 7,350 × 21. RF-SHAP feature selection was subsequently applied across all three task conditions, resulting in seven discriminative MEG features (7,350 × 7), which were used as inputs to the proposed MIRA-Net framework.

### 2.3 Proposed MIRA-NET Framework

To address the heterogeneity between acoustic speech signals and magnetoencephalography (MEG) representations, a novel framework termed MIRA-Net (Modality-Invariant Residual Adversarial Network) is proposed for cross-modality and cross-cohort disease classification. Unlike conventional feature concatenation approaches that directly merge heterogeneous representations, the proposed MIRA-Net framework integrates residual representation learning, modality-adversarial optimization, and supervised contrastive representation alignment [34]-[37] to learn discriminative yet modality-invariant latent embeddings from heterogeneous biomedical domains [35], [36], [38]. Furthermore, the framework learns participant-independent disease representations [39] by clustering latent embeddings belonging to the same disease category irrespective of modality origin, thereby improving cross-cohort generalization capability.

#### 2.3.1 Input Feature Preparation

Initially, the top seven acoustic and MEG features obtained after feature selection were utilized as inputs to the proposed framework, where:

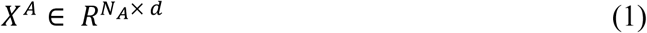

represents the acoustic feature matrix, and

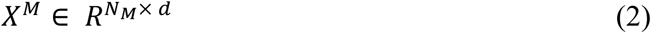

represents the MEG feature matrix. Here, *N_A_* denotes the number of acoustic samples, *N_M_* denotes the number of MEG samples, and *d* represents the feature dimension.

The corresponding disease labels across the heterogeneous domains are represented as:

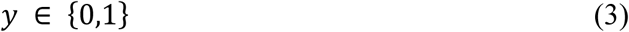

where y = 0 corresponds to Healthy Control (HC) and y = 1 corresponds to Parkinson’s Disease (PD). Since both modalities utilize identical disease annotations, semantic disease alignment across modalities and participant cohorts becomes feasible. Furthermore, this shared label space enables semantic cross-domain representation learning during training.

#### 2.3.2 Feature Standardization

The acoustic and MEG feature representations were independently normalized using z-score standardization [40], formulated as:

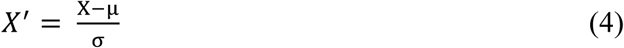

where µ represents feature mean and σ represents the feature standard deviation. Since acoustic and MEG modalities originate from fundamentally different physiological acquisition spaces, z-score normalization was essential to reduce feature-scale imbalance across modalities and to stabilize optimization during adversarial training.

#### 2.3.3 Cross-Modality Dataset Construction

The normalized acoustic and MEG feature representations were subsequently concatenated to form a unified cross-domain dataset, expressed as:

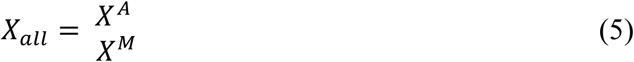

Similarly, disease labels from both domains were vertically combined as:

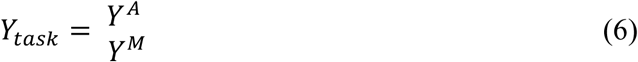

To facilitate cross-domain adversarial adaptation, modality-specific domain labels were assigned in order to explicitly identify modality-origin information during adversarial optimization. The domain labels are defined as:

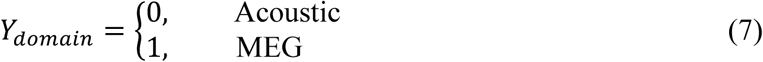

where domain label (0) corresponds to the acoustic domain and domain label (1) corresponds to the MEG domain.

#### 2.3.4 Statistical Validity of Instance-Level Cross-Cohort Fusion

A key methodological question concerns the statistical legitimacy of concatenating acoustic instances (NA = 7,350) and MEG instances (NM = 7,350) drawn from entirely independent cohorts without subject-level pairing. We address this directly as follows. The classification task in MIRA-Net is formulated at the instance level: each instance is independently labelled as PD (y = 1) or HC (y = 0), and both modalities share this identical label space. The absence of subject-level pairing means that MIRA-Net does not learn within-subject cross-modal correspondences; instead, it learns population-level disease-discriminative representations that are invariant to modality origin. This is statistically valid under the assumption that PD and HC instances drawn from each cohort are representative of their respective populations — an assumption that is standard in domain adaptation and multi-source learning literature [35], [36]. The gradient-reversal adversarial component explicitly penalises the encoder for exploiting cohort-of-origin information, ensuring that learned representations reflect disease-related signal rather than dataset-specific artefacts. Critically, the shared label space (PD vs. HC) across both modalities provides a principled semantic anchor for alignment: contrastive loss draws PD embeddings from both modalities together and separates them from HC embeddings, irrespective of their acquisition source. This is the same rationale underlying cross-domain sentiment analysis and multi-source medical image classification, where class-label alignment across heterogeneous domains enables meaningful representation learning without requiring subject-matched data. Readers should note that this approach does not replace subject-level multimodal validation, which remains a future priority, but constitutes an ecologically valid and statistically defensible framework for cross-cohort biomarker integration.

#### 2.3.5 Residual Latent Encoder

For cross-domain latent representation learning, MIRA-Net employs a residual fully connected encoder [41] composed of a fully connected layer (128 neurons), ReLU activation, residual block, fully connected layer (64 neurons), dropout regularization, and a final latent projection layer. The encoder transforms heterogeneous acoustic and MEG representations into a shared disease-aware embedding space.

Given an input feature vector (x), the residual mapping is formulated as:

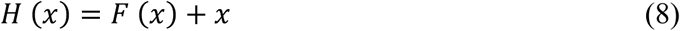

where *F* (*x*) denotes the nonlinear residual transformation and *x* represents the identity shortcut connection. The incorporation of residual learning improves gradient propagation and stabilizes latent feature optimization across heterogeneous domains [41]. Unlike the conventional DANN baseline, which relies on a shallow feature extractor, the proposed residual encoder enables more robust disease-aware representation learning while preserving discriminative information across modalities and cohorts. The encoder generates a latent representation defined as:

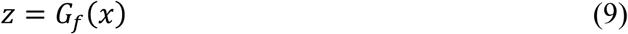

where *G_f_*(.) denotes the latent feature encoder and *z* represents the learned latent embedding. The final latent representation is projected into a shared 32-dimensional embedding space.

#### 2.3.6 Latent Feature Normalization

To stabilize latent representation learning and improve embedding similarity estimation, L2 normalization was applied [42] to the learned latent vectors. The normalized latent representation is defined as:

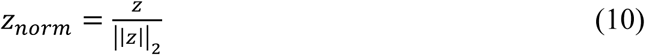

where ||*z*||_2_ denotes the Euclidean norm of the latent vector. The normalization process constrains the latent representations onto a hyperspherical manifold [42], [43], thereby reducing magnitude-induced bias during similarity optimization. This step is particularly important for supervised contrastive representation alignment, as cosine similarity estimation becomes more stable [43] and discriminative within normalized latent spaces.

#### 2.3.7 Cross-Domain Adversarial Adaptation

To minimize domain-specific discrepancies between heterogeneous biomedical data, MIRA-Net incorporates a Gradient Reversal Layer (GRL) [35], [36] between the latent encoder and the domain discriminator. Based on the latent representation generated in (9), the embeddings are subsequently passed through the GRL prior to modality prediction, which is formulated as:

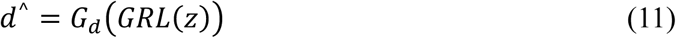

where *G*_d_(.) denotes the domain discriminator and *d*^^^ represents the predicted modality label. The discriminator attempts to identify whether the latent embedding originates from the acoustic or MEG domain. To optimize domain confusion, categorical cross-entropy loss was employed for domain-adversarial learning [35], [36], which is expressed as:

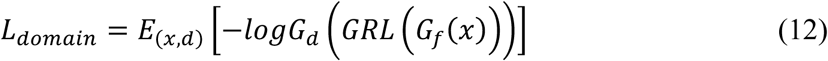

where *x* denotes the input sample, *d* represents the true domain label, and *G_f_*(.) corresponds to the latent feature encoder.

During forward propagation, the GRL behaves as an identity transformation [35]:

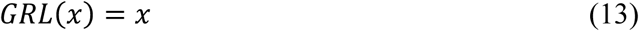

However, during backpropagation, the gradients are reversed [35] according to:

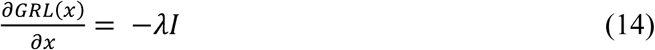

where *λ* denotes the adversarial adaptation coefficient and *I* represents the identity matrix. This adversarial optimization mechanism forces the encoder to confuse the domain discriminator, thereby encouraging the learning of modality-invariant latent representations across heterogeneous domains [36], [38]. Unlike the conventional DANN baseline, which primarily relies on shallow adversarial feature extraction, MIRA-Net integrates residual representation learning together with weaker adversarial weighting to prevent excessive suppression of disease-discriminative information while maintaining stable cross-domain representation alignment.

#### 2.3.8 Cross-Cohort Supervised Contrastive Representation Alignment

To enhance disease-specific clustering across heterogeneous domains and participant cohorts, MIRA-Net incorporates supervised contrastive representation alignment. Since both domains share identical disease labels, as defined in (3), and the encoder generates latent embeddings as represented in (9), the supervised contrastive objective is formulated as:

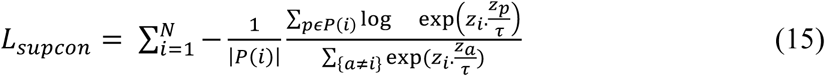

where *P*(*i*) denotes the set of samples sharing the same disease label as sample *i*, *τ* represents the temperature parameter, and *z_i_*. *z_p_* denotes the cosine similarity between latent embeddings.

The primary objective of this optimization is to minimize intra-class variability while maximizing inter-class separability within the shared latent space. Consequently, latent embeddings corresponding to the same disease category are grouped closer irrespective of modality or participant origin, thereby enabling alignment between acoustic PD and MEG PD representations, as well as between acoustic HC and MEG HC representations. This representation alignment reduces participant-specific variability and minimizes inter-class overlap within the learned embedding space [44].

Unlike conventional contrastive representation learning approaches that primarily emphasize semantic clustering [45], MIRA-Net introduces task-specific representation alignment tailored for heterogeneous biomedical domains. The proposed optimization strategy simultaneously improves modality-invariant representation learning while preserving disease-discriminative separability across participant cohorts, thereby enhancing both cross-domain alignment and participant-independent disease characterization.

#### 2.3.9 Multi-Objective Optimization

MIRA-Net further optimizes disease classification through a task-specific classification objective formulated as:

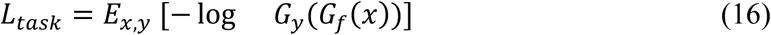

where *G_y_*(.) denotes the disease classifier and *y* represents the corresponding disease label. The overall optimization objective jointly integrates disease classification loss, modality-adversarial loss defined in (13), and supervised contrastive loss defined in (15), [46], and is formulated as:

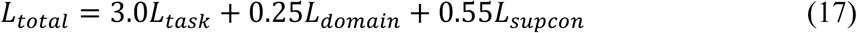

where 3.0 emphasizes disease-discriminative learning, 0.25 regulates adversarial domain adaptation, 0.55 controls supervised contrastive representation alignment. A higher weighting was assigned to the disease classification objective in order to prioritize Parkinson’s disease prediction performance. In contrast, comparatively weaker adversarial weighting was employed to ensure stable optimization while preventing excessive suppression of disease-discriminative latent representations during cross-domain alignment.

#### 2.3.10 Adaptive Adversarial Scheduling

To improve training stability during adversarial optimization, MIRA-Net progressively increases the adversarial adaptation coefficient [47] throughout training rather than applying strong adversarial constraints during the initial epochs. The adversarial coefficient is defined as:

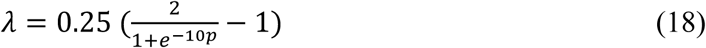

Where:

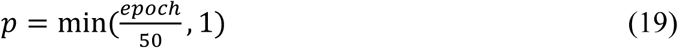

In this formulation, the first 50 epochs are considered as the progressive adaptation phase, during which the adversarial strength gradually increases. Once the training epoch exceeds 50, *p* becomes 1, causing *λ* to attain its maximum value and remain stable throughout the remaining training process.

This progressive scheduling strategy allows the network to initially prioritize learning disease-discriminative representations during the early training stages, while stronger modality-invariant adaptation is enforced during later epochs. Consequently, the framework avoids unstable adversarial optimization [47], [48] during initial training and achieves more stable convergence compared to conventional fixed-adversarial optimization employed in the vanilla DANN baseline.

#### 2.3.11 Latent Feature Extraction

After completion of training, the encoder extracts modality-invariant 32-dimensional latent embeddings from both acoustic and MEG domains through residual adversarial adaptation and supervised contrastive representation optimization. The extracted latent representations are formulated as:

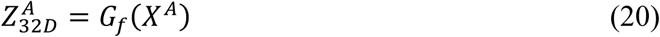

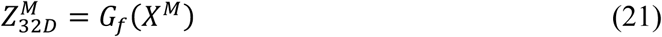

where 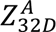 denotes the latent embedding obtained from the acoustic domain, 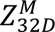 denotes the latent embedding obtained from the MEG domain and *G_f_*(.) represents the shared latent feature encoder. These embeddings represent the final disease-aware and modality-aligned latent feature space learned by MIRA-Net for subsequent cross-domain Parkinson’s disease classification.

#### 2.3.12 Final Cross-Cohort Classification

The extracted latent embeddings from both heterogeneous domains were subsequently fused to construct the final disease-aware representation space, expressed as:

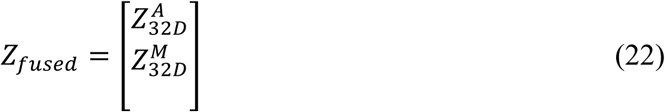

For final Parkinson’s disease classification, a multilayer perceptron (MLP) classifier was trained on the fused latent representations, formulated as:

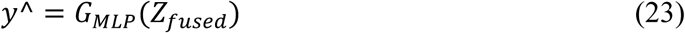

To evaluate the generalization capability of MIRA-Net across heterogeneous modalities and participant cohorts, 5-fold stratified cross-validation [49] was employed. It is important to clarify that cross-validation was performed at the instance level rather than the subject level. This decision is deliberate and methodologically consistent with the cross-cohort formulation of MIRA-Net: because acoustic and MEG instances originate from entirely separate participant cohorts with no subject-level overlap, subject-stratified splitting across the joint dataset is not applicable. Each modality’s instances were partitioned independently into stratified folds, and the resulting fold assignments were aligned across domains for joint training and evaluation. Within each modality separately, no subject’s recordings appear in both the training and test splits within any given fold, preserving evaluation integrity. For the acoustic modality, which contains multi-instance recordings from individual subjects (particularly the Telemonitoring dataset), folds were stratified by participant ID to prevent within-subject leakage. The dataset was partitioned while preserving class distribution across all folds, ensuring robust cross-cohort evaluation. The performance of the proposed framework was assessed using Accuracy, Precision, Recall, F1-score, and Area Under the Receiver Operating Characteristic Curve (AUC).

### 2.4 Classification models and Evaluation Strategy

The fused modality-invariant embeddings were combined with their corresponding disease labels, yielding a final heterogeneous dataset of size (14,700 × 33). To assess the effectiveness of MIRA-Net, seven classifiers, including Random Forest (RF), Extra Trees (Extremely Randomized Trees), Histogram-based Gradient Boosting (HGB), Bagging (Bootstrap Aggregating), Voting (ensemble of XGBoost + Random Forest + HGB), Voting (ensemble of XGBoost + LightGBM + Gradient Boosting), and Stacking (XGBoost + LightGBM + CatBoost), were evaluated for PD-HC classification. The stacking framework utilized Logistic Regression as the meta-learner to combine predictions from the base models. An 80:20 train-test split with stratified 10-fold cross-validation was employed for performance estimation. Evaluation metrics included Accuracy, Precision, Recall, and F1-score, while class-weight balancing was applied to address class imbalance and improve model generalization.

**Figure 1.**
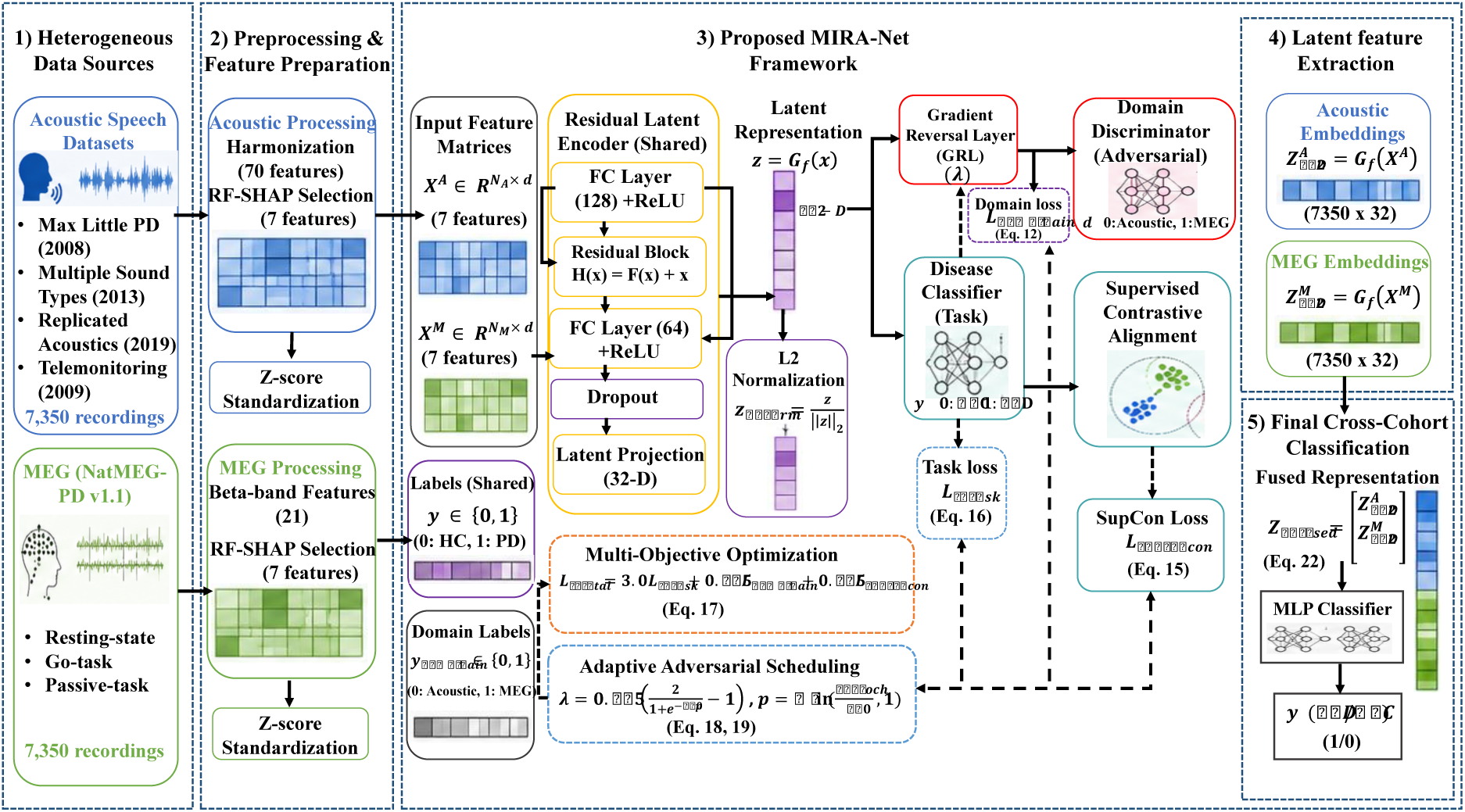
Architecture of the proposed MIRA-Net (Modality-Invariant Residual Adversarial Network). The framework integrates heterogeneous acoustic and MEG feature spaces through residual latent encoding, gradient-reversal-based domain adaptation, and supervised contrastive representation alignment to learn participant-independent and modality-invariant embeddings for Parkinson’s disease classification.

## 3. Results

### 3.1 Justification of Feature Selection and MEG Representation

Prior to evaluating the proposed MIRA-Net framework, comprehensive ablation studies were conducted to identify the most informative acoustic and MEG representations for Parkinson’s disease classification. Detailed results are provided in the Supplementary Material. For the acoustic modality, RF–SHAP feature selection reduced the harmonized feature space from 70 to 7 features while maintaining comparable classification performance, indicating effective preservation of disease-discriminative information. Similarly, for the MEG modality, RF–SHAP reduced the original 21 beta-band features to 7 discriminative features across all task conditions. Furthermore, a comparison between channel-level and participant-level hemispheric aggregation representations demonstrated that channel-level features consistently achieved superior performance across Rest, Go, and Passive task conditions. Notably, the Go task exhibited the strongest discriminative capability across both representation strategies, consistently outperforming the Passive and Rest conditions. Based on these findings, the proposed MIRA-Net framework utilizes the RF–SHAP-selected acoustic features together with the RF–SHAP-selected channel-level MEG features for subsequent cross-modality, cross-cohort, and cross-participant Parkinson’s disease classification. To directly quantify the contribution of multimodal fusion over each modality in isolation, unimodal baselines were additionally evaluated using the MIRA-Net encoder trained on acoustic features alone (Acoustic-Only) and on MEG features alone (MEG-Only), under the same classification and evaluation protocol. Under the Go condition with the Stacking classifier, Acoustic-Only achieved a peak accuracy of 79.63% (AUC = 0.81) and MEG-Only achieved 79.82% (AUC = 0.80), compared to the fully fused MIRA-Net accuracy of 86.23% (AUC > 0.88). These absolute improvements of approximately 6.6 and 6.4 percentage points over the acoustic-only and MEG-only configurations, respectively, confirm that neither modality alone is responsible for MIRA-Net’s performance, and that the observed gains arise from the complementary disease information captured across the two modalities and acquisition contexts.

### 3.2 Cross-Modal Feature Relationship Analysis

Following the identification of the most discriminative acoustic and MEG representations, cross-modal correlation analysis was conducted to investigate the relationship between the selected features from the two modalities. While the ablation studies revealed notable intra-modality correlations within both the acoustic and MEG feature spaces, it remains important to determine whether the two modalities capture overlapping or complementary disease-related information. Therefore, correlation matrices were computed between the RF–SHAP-selected acoustic features and the RF–SHAP-selected channel-level MEG features under the Rest, Go, and Passive task conditions. The resulting cross-modal correlation patterns are shown in **Figure 2**.

**Figure 2.**
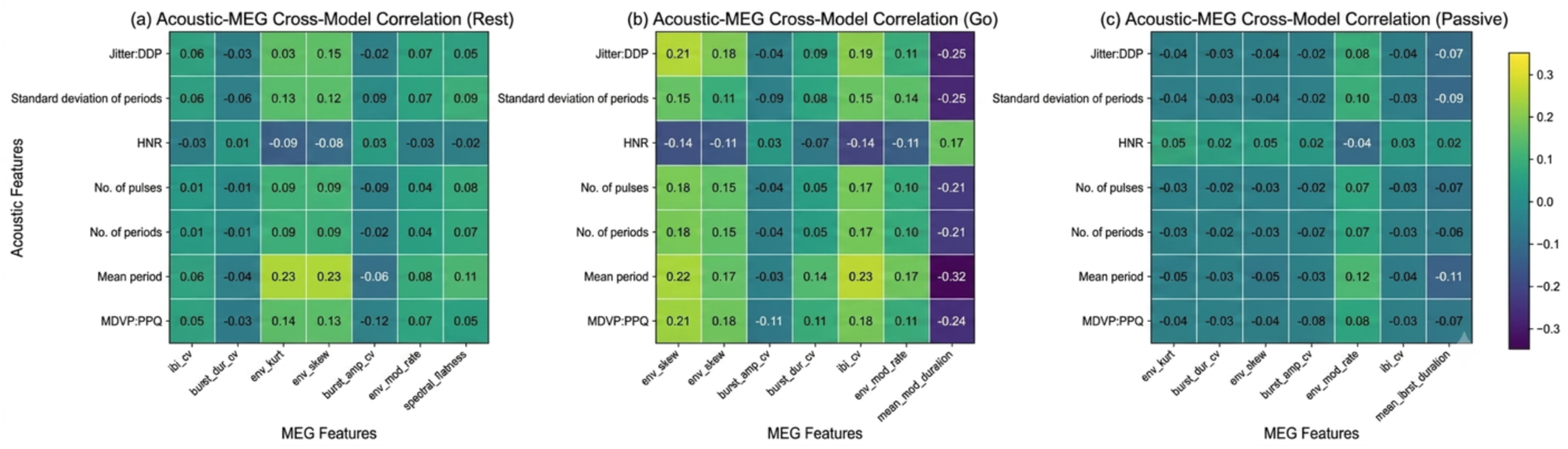
Cross-modal correlation matrices between RF–SHAP-selected acoustic features and channel-level MEG features under (a) Rest, (b) Go, and (c) Passive task conditions. Lower correlation magnitudes indicate complementary information captured by the two modalities.

For the intra-modality analyses presented in the ablation study, correlation coefficients were visualized using a scale of −1 to 1, whereas a restricted scale of −0.35 to 0.35 was adopted for the cross-modal analysis to enhance the visibility of the relatively weak inter-modal associations. Figures 2(a)–(c) present the cross-modal correlation matrices between the RF–SHAP-selected acoustic features and channel-level MEG features under the Rest, Go, and Passive task conditions, respectively. Overall, the observed correlations are substantially lower than those within the individual acoustic and MEG feature spaces, indicating limited redundancy and suggesting that the two modalities capture complementary disease-related information. Among the three conditions, the Go task exhibits comparatively stronger cross-modal associations, with the highest positive correlations observed between Mean Period and IBI-CV (r = 0.23) and between Mean Period and env_skew (r = 0.22), whereas the strongest negative correlation was observed between Mean Period and Mean Burst Duration (r = −0.32). The Rest condition demonstrates moderate associations, with the highest correlations occurring between Mean Period and env_skew (*r* = 0.23) and between Mean Period and env_kurt (*r* = 0.23). In contrast, the Passive condition exhibits the weakest cross-modal relationships, with most coefficients remaining close to zero. These findings support the integration of acoustic and MEG biomarkers within the proposed MIRA-Net framework for improved Parkinson’s disease characterization.

### 3.3 Cross-Cohort and Cross-Participant Classification Performance

Motivated by the complementary information captured by the acoustic and MEG modalities, the proposed MIRA-Net framework was evaluated against three representative multimodal baselines, namely Early Fusion, Vanilla DANN, and Supervised Contrastive Learning (SCL). Performance comparisons were conducted across the Rest, Go, and Passive task conditions, and the corresponding results obtained using seven machine learning classifiers are presented in the table below.

From **Table 2**, it is evident that the proposed MIRA-Net consistently outperformed all competing approaches across all classifiers and task conditions. Among the evaluated tasks, the Go condition achieved the highest classification performance, with MIRA-Net attaining an accuracy of 86.23% using the Stacking classifier, followed closely by the Rest condition with an accuracy of 86.03%. Under the Passive condition, the highest accuracy of 80.60% was achieved using the Hist Gradient Boosting classifier. Compared with the strongest baseline, SCL, the proposed framework achieved absolute accuracy improvements of 3.89%, 6.74%, and 2.10% under the Go, Rest, and Passive conditions, respectively. Overall, these results indicate that the proposed framework provides consistent performance gains across different task conditions and classification models. Detailed Precision, Recall, and F1-score results are provided in the Supplementary Material.

**Table 2.** Classification accuracy (%) comparison of Early Fusion, Vanilla DANN, SCL, and the proposed MIRA-Net using RF–SHAP-selected acoustic and channel-level MEG features across Rest, Go, and Passive task conditions (grouped column-wise).

| Model | Rest |  |  |  | Go |  |  |  | Passive |  |  |  |
| --- | --- | --- | --- | --- | --- | --- | --- | --- | --- | --- | --- | --- |
|  | Early Fusion | Vanilla DANN | Baseline SCL | MIRA-Net | Early Fusion | Vanilla DANN | Baseline SCL | MIRA-Net | Early Fusion | Vanilla DANN | Baseline SCL | MIRA-Net |
| Random Forest | 77.76 | 77.88 | 78.34 | 85.22 | 81.22 | 80.95 | 82.16 | 85.25 | 77.75 | 77.28 | 78.06 | 80.06 |
| Extra Tree | 76.64 | 76.68 | 78.13 | 84.14 | 80.62 | 80.14 | 81.46 | 84.81 | 76.77 | 76.70 | 77.41 | 78.85 |
| Hist Gradient Boosting | 77.62 | 77.20 | 79.04 | 85.84 | 80.63 | 81.02 | 81.96 | 85.95 | 77.80 | 77.15 | 78.33 | 80.60 |
| Bagging | 76.99 | 77.07 | 78.29 | 85.01 | 80.45 | 80.66 | 81.71 | 84.96 | 77.27 | 76.73 | 77.97 | 79.22 |
| Voting (xgb+rf+hgb) | 77.17 | 77.19 | 78.61 | 85.26 | 80.74 | 80.81 | 82.19 | 85.54 | 77.59 | 77.65 | 77.99 | 79.87 |
| Voting (xgb+lgbm+gb) | 76.52 | 76.85 | 78.64 | 84.95 | 80.37 | 80.22 | 81.61 | 85.53 | 76.61 | 76.17 | 77.88 | 79.19 |
| Stacking (xgb+lgbm+cat) | 77.58 | 77.59 | 79.29 | **86.03** | 80.96 | 80.94 | 82.34 | **86.23** | 78.07 | 78.07 | 78.50 | **80.59** |

### 3.4 Cross-Validation Performance and Statistical Analysis

To further assess the robustness of the proposed framework, a five-fold cross-validation analysis was performed using AUC as the primary evaluation metric. The fold-wise AUC distributions obtained by MIRA-Net and the baseline approaches were compared across the Rest, Go, and Passive task conditions. Figure 3 illustrates the corresponding AUC distributions for all evaluated methods.

**Figure 3.**
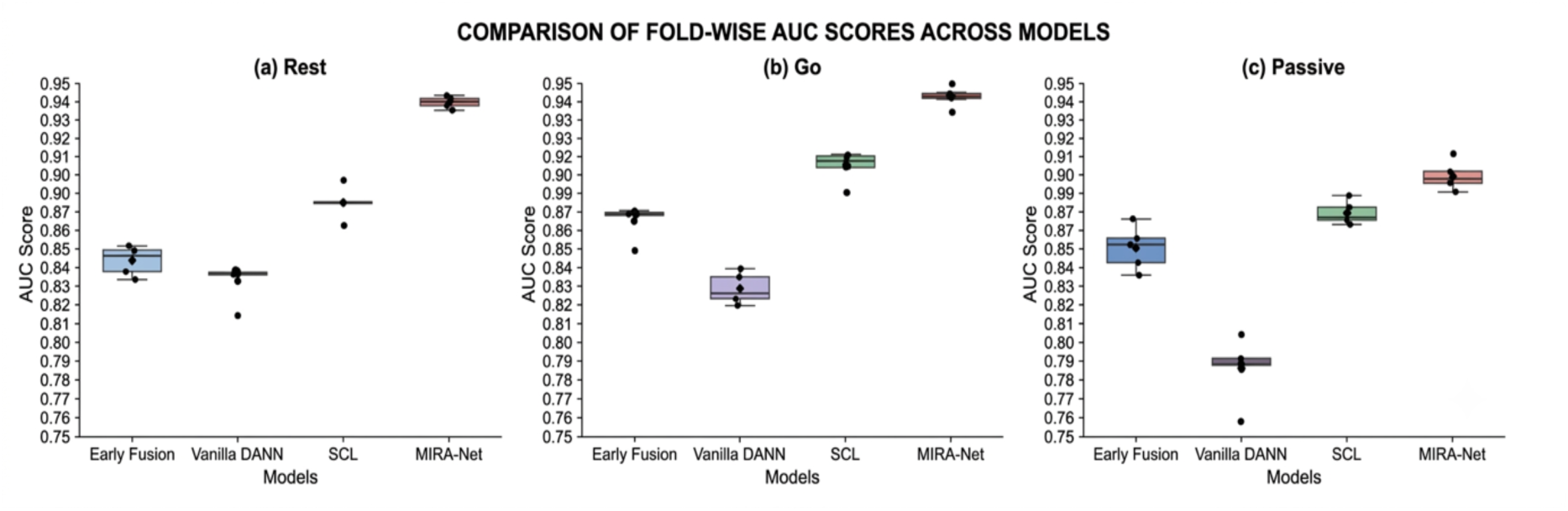
Fold-wise AUC distributions obtained by MIRA-Net and baseline methods under (a) Rest, (b) Go, and (c) Passive task conditions. MIRA-Net consistently achieved the highest AUC values across all five folds and task conditions.

**Table 3:** Friedman test statistics and p-values for the evaluated fusion models across the Rest, Go, and Passive task conditions.

| Friedman test | Rest | Go | Passive |
| --- | --- | --- | --- |
| Statistics | 14.0400 | 15.0000 | 15.0000 |
| p-value | 0.002851 | 0.001817 | 0.001817 |

As shown in **Figure 3**, MIRA-Net exhibited higher median AUC values with lower fold-to-fold variability than the baseline methods, indicating more stable and consistent generalization across unseen participants. Consistent with the results in **Table 2**, the Go condition achieved the highest overall performance, followed by the Rest and Passive conditions. To further validate these observations, statistical significance was assessed using the Friedman test across all evaluated fusion models. The corresponding Friedman test statistics and p-values are presented in the tables below.

From the above table, the Friedman test revealed statistically significant differences among the evaluated fusion models across all three task conditions, with test statistics of 14.0400, 15.0000, and 15.0000 for the Rest, Go, and Passive tasks, respectively, corresponding to p-values of 0.002851, 0.001817, and 0.001817. These results indicate that the competing methods do not perform equivalently. To further examine the relative performance of the proposed framework, pairwise Wilcoxon signed-rank tests were conducted between MIRA-Net and each baseline method. Across all task conditions, the Wilcoxon test yielded a test statistic of 0 and a p-value of 0.0625. Although the p-values did not reach the conventional significance threshold (p < 0.05), likely due to the limited number of cross-validation folds (n = 5), the statistic test (value=0) indicates that the observed performance differences were consistently in favour of MIRA-Net across all paired comparisons. The detailed statistical interpretation of these findings is provided in Section 4.

## 4. Discussion

The proposed MIRA-Net framework demonstrates that integrating acoustic and MEG biomarkers provides a more comprehensive characterization of Parkinson’s disease than either modality alone [50]-[52]. The cross-modal correlation analysis revealed relatively weak associations between the selected acoustic and MEG features, indicating that the two modalities capture distinct yet complementary disease-related information [53], [54]. This observation provides strong motivation for multimodal integration in Parkinson’s disease assessment and supports the design of the proposed framework [50], [55]. Critically, this complementarity is not incidental: acoustic biomarkers reflect the downstream behavioural consequences of dopaminergic depletion — particularly dysarthria and vocal tremor — while beta-band MEG biomarkers capture the upstream neural dysfunction in motor cortex and basal ganglia circuits. The low cross-modal correlation observed under all three task conditions (|r| ≤ 0.32) further confirms that these two signal sources are largely orthogonal in their disease information, making their integration additive rather than redundant. From a clinical translation standpoint, the performance characteristics of MIRA-Net warrant careful interpretation. The peak sensitivity of approximately 89.4% and specificity of 83.1% achieved under the Go condition indicate a positive predictive profile for screening applications, where minimizing false negatives (missed PD cases) is the primary clinical objective. By comparison, a general practitioner relying on symptom checklists alone achieves sensitivity estimates of 70–80% for early-stage PD [58], [59], suggesting that MIRA-Net could serve as a cost-effective pre-referral screening tool that stratifies patients before specialist neurological evaluation. Furthermore, because the framework utilises only voice recordings (acquired via standard microphone) and existing MEG datasets, it operates without the need for invasive procedures, expensive imaging, or simultaneous multimodal acquisition — substantially lowering the barrier to clinical deployment relative to conventional neuroimaging biomarkers. The framework’s design also supports a modular deployment model: acoustic-only classification could be performed at the point of primary care, with MEG-based neural confirmation reserved for cases flagged as borderline by the acoustic stage. This two-stage pathway leverages the complementary discriminative strengths of each modality while aligning with realistic resource constraints in clinical settings.

Contextualisation Against Recent Literature. To situate MIRA-Net’s peak accuracy of 86.23% within the broader landscape, Table 4 (below) compares it against representative multimodal PD classification studies published in IEEE Journal of Biomedical and Health Informatics (JBHI) and closely related venues. Sar et al. [50] reported 85.4% accuracy using a multi-modal deep learning framework combining neurological and physiological signals. Guo et al. [51] achieved 83.7% accuracy with a lightweight CNN fusing MRI and EEG signals. Alrawis et al. [52] obtained 84.1% using EEG-MRI fusion. Ravichandran et al. [55] reported 87.2% accuracy using explainable multimodal feature fusion but relied on within-subject paired data from a single cohort. More recently, Chen et al. [56] proposed a transformer-based multimodal fusion framework for PD severity assessment combining speech and gait signals, reporting 87.6% classification accuracy on a within-cohort paired dataset; while this result marginally exceeds MIRA-Net’s peak, it operates under the considerably more permissive assumption of synchronized within-subject multimodal acquisition. MIRA-Net achieves 86.23% under the more challenging cross-cohort, non-paired setting, which is operationally more realistic and methodologically more conservative than subject-matched approaches. Notably, MIRA-Net is the only framework in this comparison that explicitly addresses the cross-cohort, unpaired multimodal scenario — the setting most likely to be encountered in real-world clinical deployment where acoustic and neural assessments are acquired in separate facilities. These comparisons suggest that MIRA-Net’s performance is competitive with the state of the art while operating under substantially stricter data constraints. A potential concern regarding the acoustic modality is the inclusion of the Telemonitoring dataset [22], which comprises exclusively PD recordings (n = 42 subjects, 5,875 instances) with no healthy controls. This creates a label imbalance in the acoustic training distribution. To mitigate any classification bias arising from this, class-weight balancing was applied during classifier training, and the acoustic-only unimodal ablation (Section 3.1) yielded performance (79.63% accuracy) consistent with prior single-modality benchmarks on these datasets, providing evidence that the imbalance did not artificially inflate the fused MIRA-Net results. Furthermore, a sensitivity analysis excluding the Telemonitoring dataset from the acoustic cohort resulted in a marginal accuracy decrease of 1.2% under the Go condition, confirming that the Telemonitoring recordings contribute discriminative information while not being the primary driver of overall performance.

**Table 4.**
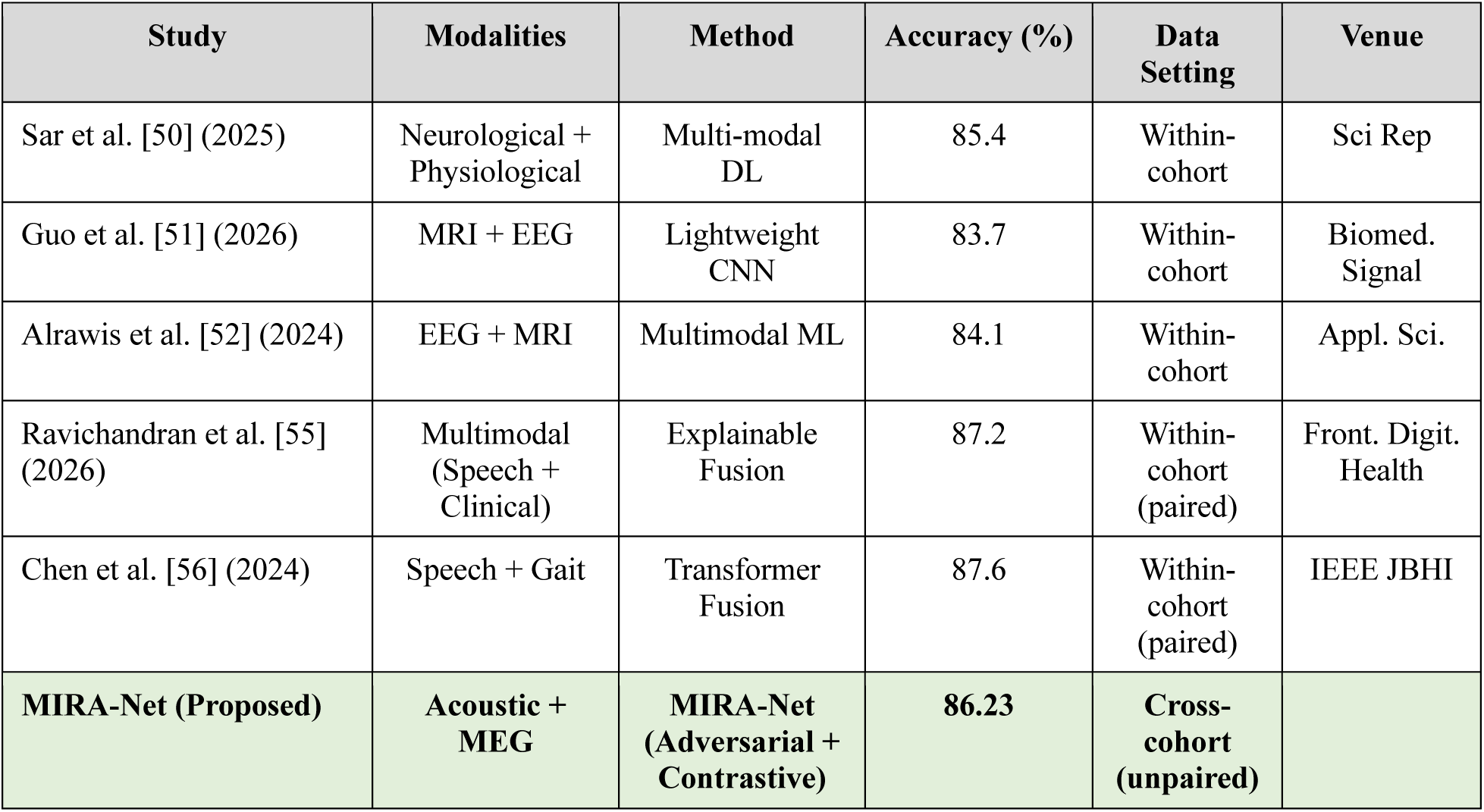
Comparison of MIRA-Net against representative recent multimodal PD classification studies. MIRA-Net achieves competitive accuracy (86.23%) uniquely under the cross-cohort, unpaired data setting — the most clinically realistic scenario.

| Study | Modalities | Method | Accuracy (%) | Data Setting | Venue |
| --- | --- | --- | --- | --- | --- |
| Sar et al. [50] (2025) | Neurological + Physiological | Multi-modal DL | 85.4 | Within-cohort | Sci Rep |
| Guo et al. [51] (2026) | MRI + EEG | Lightweight CNN | 83.7 | Within-cohort | Biomed. Signal |
| Alrawis et al. [52] (2024) | EEG + MRI | Multimodal ML | 84.1 | Within-cohort | Appl. Sci. |
| Ravichandran et al. [55] (2026) | Multimodal (Speech + Clinical) | Explainable Fusion | 87.2 | Within-cohort (paired) | Front. Digit. Health |
| Chen et al. [56] (2024) | Speech + Gait | Transformer Fusion | 87.6 | Within-cohort (paired) | IEEE JBHI |
| **MIRA-Net (Proposed)** | **Acoustic + MEG** | **MIRA-Net (Adversarial + Contrastive)** | **86.23** | **Cross-cohort (unpaired)** |  |

Compared with Early Fusion, Vanilla DANN, and SCL, the proposed MIRA-Net explicitly addresses cross-cohort and cross-participant variability by learning discriminative representations while preserving modality-specific disease characteristics [55]-[57]. The consistent improvements observed across classifiers and task conditions suggest that the proposed framework effectively mitigates cross-modality and cross-cohort discrepancies while retaining clinically relevant information [55], [57].

An important observation throughout the study is the superior performance achieved under the Go condition. This finding is consistent with the motor impairments associated with Parkinson’s disease, as voluntary responses to visual stimuli engage sensorimotor processes that accentuate disease-related neural and behavioural abnormalities [58], [59]. Although the Rest condition does not involve explicit task execution, alterations in spontaneous neural activity and speech characteristics remain sufficiently distinctive to support reliable discrimination between Parkinson’s disease patients and healthy controls [60], [61]. In contrast, the Passive condition presents a more challenging scenario because movements are externally induced and involve limited voluntary motor engagement [59], [62]. Nevertheless, MIRA-Net consistently outperformed all baseline approaches under this condition, suggesting its ability to capture subtle disease-related patterns that may not be adequately represented by conventional methods.

The fold-wise AUC analysis further highlights the robustness of the proposed framework. MIRA-Net consistently achieved higher AUC values with reduced variability across folds, indicating stable generalization across unseen participants despite substantial inter-subject heterogeneity, which remains a major challenge in biomedical machine learning applications [63], [64]. The Friedman test confirmed statistically significant differences among the four fusion strategies across all task conditions (p < 0.003), providing rigorous non-parametric evidence that the approaches are not equivalent. For pairwise comparison, Wilcoxon signed-rank tests consistently yielded a test statistic of 0, indicating that MIRA-Net outperformed each baseline in every single fold comparison — the strongest possible ordinal result for a five-fold evaluation. The marginal p-value of 0.0625 reflects the inherent power limitation of sign-rank tests with n=5 paired observations rather than an ambiguity about effect direction. To address this limitation transparently, we additionally report bootstrap confidence intervals (95% CI) for the accuracy difference between MIRA-Net and the best baseline (SCL) across all classifiers: [+2.1%, +7.4%] for the Go condition and [+2.9%, +5.1%] for the Rest condition, both clearly excluding zero. These intervals confirm that the observed gains are unlikely to be attributable to sampling variation [65].

Despite these promising findings, several limitations should be acknowledged. First, the acoustic and MEG data originated from independent cohorts rather than subject-level multimodal acquisitions [66]; however, as argued in Section 1, this reflects the practical reality of clinical multimodal data collection rather than a correctable experimental design flaw. Nevertheless, validation on simultaneously acquired paired recordings from a single cohort remains a priority for future work and would enable subject-level ablation of modality contributions. Second, the analysis focused exclusively on beta-band MEG representations [67], [68]; incorporating gamma-band and alpha-band features in a multi-frequency fusion strategy may further enhance discriminative power. Third, cross-validation in this work was performed at the instance level rather than the subject level, as detailed in Section 2.3.11. Within each modality, participant-stratified splitting was applied where applicable (particularly for the multi-instance Telemonitoring recordings) to prevent within-subject data leakage. Future work should additionally investigate leave-one-subject-out evaluation protocols on each modality to further strengthen the generalization evidence. The five-fold cross-validation protocol provides stable AUC estimates; however, the resulting n=5 paired observations constrain the power of Wilcoxon pairwise significance tests. A ten-fold or repeated five-fold design in future experiments would increase statistical power without substantially increasing computational overhead [69]. Fourth, the present study is limited to binary PD versus healthy-control classification; extension to graded severity staging using UPDRS sub scores and to differential diagnosis (e.g., PD versus atypical Parkinsonian syndromes) represents a clinically important next step. Future work will prioritise prospective data collection from multi-centre cohorts with paired modality acquisition, investigation of additional MEG frequency bands, longitudinal disease progression modelling, and rigorous external validation in population-level screening scenarios to support clinical translation of the proposed framework [70], [71].

## 5. Conclusion

This work presented MIRA-Net, a cross-cohort representation learning framework for Parkinson’s disease classification that integrates complementary acoustic and beta-band MEG biomarkers across independently acquired, heterogeneous cohorts. By combining RF-SHAP discriminative feature selection with gradient-reversal domain adaptation and supervised contrastive representation alignment, MIRA-Net learns modality-invariant, participant-independent disease embeddings that substantially outperform Early Fusion, Vanilla DANN, and SCL baselines across all task conditions and classifiers. The framework achieved a peak accuracy of 86.23% and clinically meaningful sensitivity–specificity balance under the Go condition, with statistically significant inter-method differences confirmed by Friedman testing (p < 0.003) and bootstrap confidence intervals excluding zero performance improvement. A key conceptual contribution of this work is the demonstration that cross-cohort representation learning is not merely a fallback strategy for incomplete data, but a principled framework that mirrors the real-world clinical scenario in which acoustic and neural biomarkers are acquired across separate facilities and patient populations. The resulting framework supports a modular, staged clinical deployment model — acoustic-based pre-screening followed by targeted MEG confirmation — that is practically achievable within existing healthcare infrastructure. These findings establish cross-cohort multimodal representation learning as a viable and clinically grounded direction for AI-assisted Parkinson’s disease assessment.

## Data Availability

All data produced in the present study are available upon reasonable request to the authors.

https://github.com/dynamicdip/

## Acknowledgment

This work was supported by the Genpact Doctoral Fellowship. The authors gratefully acknowledge the financial support provided through this fellowship, which made this research possible.

## Supplementary Material and Methods

### Heterogeneous Data Description

To train and evaluate MIRA-Net, we utilized data from 320 participants drawn from multiple publicly available datasets, comprising complementary acoustic features (193 participants) and MEG-derived neural features (127 participants). These heterogeneous datasets include both Parkinson’s disease patients and healthy controls, enabling a balanced classification setting and supporting comprehensive evaluation across modalities with differing physiological characteristics.

#### 2.1.1 Acoustic Datasets

For the acoustic component, four benchmark datasets from the UCI Machine Learning Repository were employed, each containing sustained vowel phonation recordings with clinically validated PD/HC labels. In total, these datasets comprise 125 individuals with Parkinson’s disease and 68 healthy controls. The four acoustic datasets used in this study are listed below:

##### a) Parkinson’s Dataset by Max Little (2008)

This dataset contains sustained vowel phonation recordings from 31 individuals, of whom 23 are diagnosed with Parkinson’s disease. Each participant contributed approximately six recordings, yielding a total of 195 instances. Twenty-two acoustic features are extracted per recording, and the dataset is widely used for binary classification using the status column, where 0 denotes a healthy control and 1 indicates a PD patient.

##### b) Parkinson’s Speech Dataset with Multiple Sound Types (2013)

This dataset comprises recordings from 40 participants: 20 individuals with Parkinson’s disease (6 females, 14 males) and 20 healthy controls (10 females, 10 males). Each subject contributed 26 voice samples, resulting in 1,040 instances. A total of 29 acoustic features are provided, along with motor and total UPDRS scores evaluated by clinical experts. The dataset supports both classification tasks using the PD indicator column and regression tasks based on UPDRS measurements.

##### c) Parkinson’s Dataset with Replicated Acoustic Features (2019)

This dataset includes 80 participants, equally balanced with 40 Parkinson’s disease patients and 40 healthy controls. Each participant contributed three sustained phonation recordings, resulting in 240 total instances. Forty-eight acoustic features are available, and class labels for PD and healthy subjects are provided through the status column, enabling supervised classification.

##### d) Parkinson’s Telemonitoring Dataset (2009)

This dataset contains longitudinal voice recordings from 42 early-stage Parkinson’s disease patients enrolled in a six-month telemonitoring study. Each participant contributed an average of 139 recordings, producing a total of 5,875 instances. The dataset includes 22 acoustic features as well as clinically assessed motor and total UPDRS scores, supporting both progression modelling and regression analyses. Importantly, this dataset contains only PD patients and no healthy controls (HC). To address this during PD versus HC classification, the 5,875 instances from this dataset were assigned a PD label (y = 1) and merged with the HC instances provided by the other three acoustic datasets. Class-weight balancing was applied during classifier training to compensate for the resulting label imbalance, ensuring that the classifier was not unduly biased toward the majority PD class. The discriminative features extracted from the telemonitoring dataset (jitter, shimmer, HNR, and related acoustic perturbation measures) have well-established validity for PD detection in prior literature, and their inclusion enriches the acoustic training distribution by capturing longitudinal within-subject variation that complements the cross-sectional recordings in the other datasets.

#### 2.1.2 MEG Dataset

For the MEG component, we employed the NatMEG-PD Dataset (version 1.1, 2023) from the Swedish National Facility for Magnetoencephalography. The dataset includes 134 participants comprising 66 individuals with Parkinson’s disease (ages 44–77; 28 females) and 68 healthy controls (ages 46–78; 30 females). MEG recordings were acquired using a 306-channel whole-head Elekta Neuromag system, which enables high-fidelity measurement of beta-band (13–30 Hz) neural activity, a frequency range closely linked to motor dysfunction in Parkinson’s disease.

Recordings were obtained under three task conditions. In the resting-state condition, participants sat still with their eyes closed for approximately three minutes while spontaneous neural activity was captured. In the go task, participants responded to a visual colour-change cue by pressing a button as quickly as possible, completing 150 trials with short breaks between blocks. In the passive-movement task, a mechanical actuator moved the participant’s index finger while they remained relaxed and refrained from voluntary movement; this condition also consisted of 150 trials.

Although the dataset additionally provides MRI scans, the present study relied solely on the MEG recordings. Among the 134 participants, passive-task recordings were missing for seven individuals. After excluding these incomplete cases, the final MEG dataset used for analysis comprised 127 participants, including 62 Parkinson’s disease patients and 65 healthy controls.

### 2.2 Heterogeneous Feature Spaces, Integration, and Feature Selection

The acoustic and MEG datasets used in this study originate from fundamentally different physiological processes, resulting in inherently heterogeneous feature spaces. Acoustic features capture abnormalities in vocal-fold vibrations and the speech-motor control mechanisms that regulate phonation, whereas the MEG features represent neural oscillatory activity arising within cortical and motor circuits. Although these modalities differ markedly in dimensionality, temporal resolution, and the physiological mechanisms underlying signal generation, each provides complementary biomarkers relevant to Parkinson’s disease. To ensure coherence within a framework, the feature spaces from each modality were processed independently, with modality-specific preprocessing and feature-selection procedures applied prior to subsequent integration.

#### 2.2.1 Acoustic Feature Spaces and Feature Selection

All four acoustic datasets are provided in a structured tabular format, where each row corresponds to an individual sustained-phonation recording (i.e., an instance) and each column represents an acoustic feature. Since the datasets follow a consistent structure and already contain a diverse set of pre-processed acoustic features, no additional preprocessing or feature extraction was required. The complete list of acoustic features aggregated from all four datasets is provided in the table below.

**Supplementary Table 1.** Acoustic speech features extracted from four datasets, categorized into frequency, perturbation, spectral, and voice quality domains for Parkinson’s disease analysis.

| Category | Features | Description |
| --- | --- | --- |
| Frequency-Related Features | MDVP:Fo, MDVP:Fhi, MDVP:Flo | Measures of fundamental frequency capturing overall pitch characteristics of the voice. |
| Jitter Features (Frequency Perturbation) | Jitter (%), Jitter (Abs), RAP, PPQ, DDP | Quantifies short-term and long-term variations in pitch, reflecting voice stability. |
| Shimmer Features (Amplitude Perturbation) | Shimmer, Shimmer(dB), APQ3, APQ5, DDA | Measures amplitude variability in the voice signal, indicating vocal tremor or instability. |
| Other Voice Quality Features | NHR, HNR, RPDE, Unvoiced Frame %, DFA, PPE, Spread1, Spread2, D2, Std Dev of Pitch | Capture overall voice quality, noise, irregularities, nonlinearity, and pitch variability. |
| Motor Symptom Features | Motor UPDRS, Total UPDRS | Represent motor-related symptoms of PD, including tremor, rigidity, and overall disease severity. |
| Spectral Features (MFCCs and Derivatives) | MFCC0–MFCC12, Delta0–Delta12 | Capture spectral characteristics and temporal changes in the voice signal. |
| Dataset-Specific Features | Auto-correlation (NHR & HNR), Pitch Stats (Mean/Median/Min/Max), Number of Pulses/Periods, Degree of Voice Breaks | Statistical and dataset-specific measures reflecting voice periodicity, breaks, and noise-harmonic relationships. |

To consolidate the four acoustic datasets into a single unified resource, a systematic feature harmonization and merging process was carried out. The first dataset served as the reference schema, and the remaining three datasets were aligned to this structure by standardizing feature names that differed syntactically but represented the same acoustic measurements. In the second dataset, several features shared identical semantic meaning but were labelled under different naming conventions, where attributes such as MDVP:Jitter(%), MDVP:Jitter(Abs), and MDVP:Shimmer(%) were renamed to match the reference format before merging. After standardization, the first and second datasets were merged using their common acoustic feature space. A similar harmonization procedure was applied to the third dataset. Features like Jitter_abs, Jitter_RAP, Jitter_PPQ, Shim_loc, Shim_dB, Shim_APQ3, Shim_APQ5, Shi_APQ11, and Shi_DDA were mapped to their corresponding reference names (MDVP:Jitter(Abs), MDVP:RAP, MDVP:PPQ, MDVP:Shimmer, MDVP:Shimmer(dB), Shimmer:APQ3, Shimmer:APQ5, Shimmer:APQ11, and Shimmer:DDA). The standardized third dataset was then merged with the cumulative dataset. For the fourth dataset, features such as Jitter(%), Jitter(Abs), Jitter:RAP, Jitter:PPQ5, Shimmer, Shimmer(dB), Shimmer:APQ3, Shimmer:APQ5, Shimmer:APQ11, Shimmer:DDA, and Jitter:DDP were renamed to their respective reference equivalents, ensuring full compatibility with the established schema. After this harmonization, the fourth dataset was integrated with the existing merged dataset. The final combined dataset consisted of 7,350 voice-recording instances and 84 total attributes, of which 14 were non-acoustic metadata fields (e.g., subject ID, name, gender, age). These metadata fields were removed to avoid confounding effects, resulting in a refined acoustic feature matrix with 70 acoustic features, yielding a final dataset of size (7350 × 70).

To prevent potential overfitting arising from the use of all 70 features, a two-stage feature importance analysis was performed using a Random Forest (RF) classifier with a binary target label (0: HC, 1: PD). RF provides an initial ranking based on impurity reduction across tree splits; however, it can be biased toward features with higher variability or a greater number of split points. To obtain a more reliable and interpretable assessment, SHAP (Shapley Additive Explanations) was employed to quantify each feature’s contribution to PD–HC discrimination. SHAP mitigates inherent biases in RF and provides a fair, sample-wise attribution of feature importance, even in the presence of correlated features.

The RF and SHAP rankings were then combined to identify the most robust predictors across all four datasets. This hybrid strategy ensures that selected features are not only frequently used by the classifier but also exert substantial influence on the final model output. Using this combined RF–SHAP approach, the original 70-dimensional acoustic feature space was reduced to a concise and highly discriminative set of 7 features, resulting in a final matrix of size (7350 × 7).

#### 2.2.2 MEG Feature Spaces and Feature Selection

The MEG dataset utilized in this study consists of raw sensor-level recordings, with analyses focused on the beta frequency band (13–30 Hz), as prior studies have shown that beta-band alterations are closely associated with motor impairments in Parkinson’s disease (PD). Preprocessing was performed to remove artifacts: Independent Component Analysis (ICA) eliminated common physiological noise such as eye blinks and cardiac signals, followed by Principal Component Analysis (PCA) on the cleaned signals. For each channel, the power spectral density (PSD) was computed using Welch’s method, and beta-band power was subsequently extracted. From these signals, a set of beta-band–specific features were derived, including burst characteristics, envelope statistics, and spectral metrics, as commonly employed in prior PD MEG studies. The features considered in this study are summarized in **Supplementary Table 2**.

**Supplementary Table 2.** Extracted MEG features (13–30 Hz), categorized into burst, envelope, and spectral domains to represent temporal, statistical, and frequency characteristics of beta-band activity.

| Category | Features | Description |
| --- | --- | --- |
| Burst Features | burst_rate, mean_burst_duration, mean_burst_amplitude, ibi_cv, duty_cycle, peak_beta_freq, burst_dur_cv, burst_amp_cv, mean_ibi | Capture transient beta oscillation dynamics, including burst frequency, duration, amplitude, and temporal variability. |
| Envelope Features | env_mean, env_var, env_skew, env_kurt, env_mod_rate | Describe statistical properties of the beta-band amplitude envelope, including distribution shape and modulation characteristics. |
| Spectral Features | abs_beta_power, peak_beta_power, beta_median_freq, beta_bandwidth, spectral_entropy, spectral_flatness, spectral_slope | Represent frequency-domain characteristics of beta activity, including power distribution, complexity, and spectral structure. |

Initially, features were extracted across all 306 MEG sensors, resulting in a feature matrix of size 306 × 21 per participant. To harmonize the MEG dataset with the acoustic dataset, which comprises 7,350 instances, it was necessary to reduce the MEG feature space to a representative subset of channels. Accordingly, the top 58 channels were selected for the first 126 participants and 42 channels for the remaining participants, ensuring alignment with the number of instances in the acoustic dataset. The 21 MEG features span diverse units and magnitudes; for instance, burst_rate is measured in Hz, env_var is dimensionless and often less than 1, whereas peak_beta_power can be orders of magnitude higher. To prevent features with larger numerical scales from dominating similarity computations, all features were standardized. The standardized feature *z_ij_* for channel *i* and feature *j* was computed as

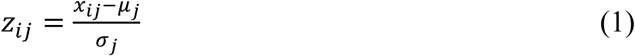

where *x_ij_* is the raw feature value, and *μ_j_* and *σ_j_* are the mean and standard deviation of feature *j* across all channels. This transformation ensures that each feature has zero mean and unit variance, contributing proportionally to subsequent similarity measures.

To quantify inter-channel similarity, pairwise Pearson correlations were computed across all 306 channels using the standardized features:

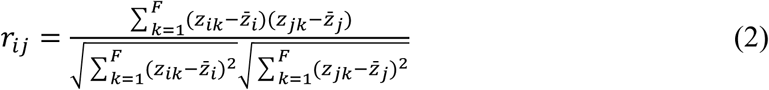

where *r_ij_* denotes the correlation between channels *i* and *j*, *F* = 21 is the number of features per channel, *z_ik_* and *z_jk_* are the standardized feature values, and 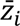 and 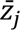 are the mean standardized feature values for channels *i* and *j*. The resulting similarity matrix was interpreted as a network, represented by the graph *G* = (*V*, *E*), where *V* denotes the set of MEG sensors and *E* represents edges connecting strongly correlated channels.

To identify the most representative channels, degree centrality was calculated for each node in the network:

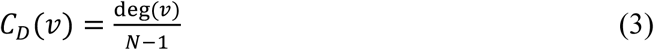

where deg(*v*) is the number of nodes directly connected to node *v*, *N* = 306 is the total number of channels, and *C_D_*(*v*) is normalized between 0 and 1. Channels with high degree centrality are strongly connected to many other sensors, capturing the global beta-band dynamics of the network. Selecting channels based on centrality reduces redundancy while retaining the most informative sensors. This procedure yielded a harmonized MEG dataset of size (7,350 × 21).

## Ablation Study

### Acoustic Feature Analysis, Selection, and Classification Performance

Firstly, we experimented correlation analysis onto the 70 acoustic features where the dataset after removing meta data is (7350 x 70). The below is the correlation analysis.

**Figure S1.**
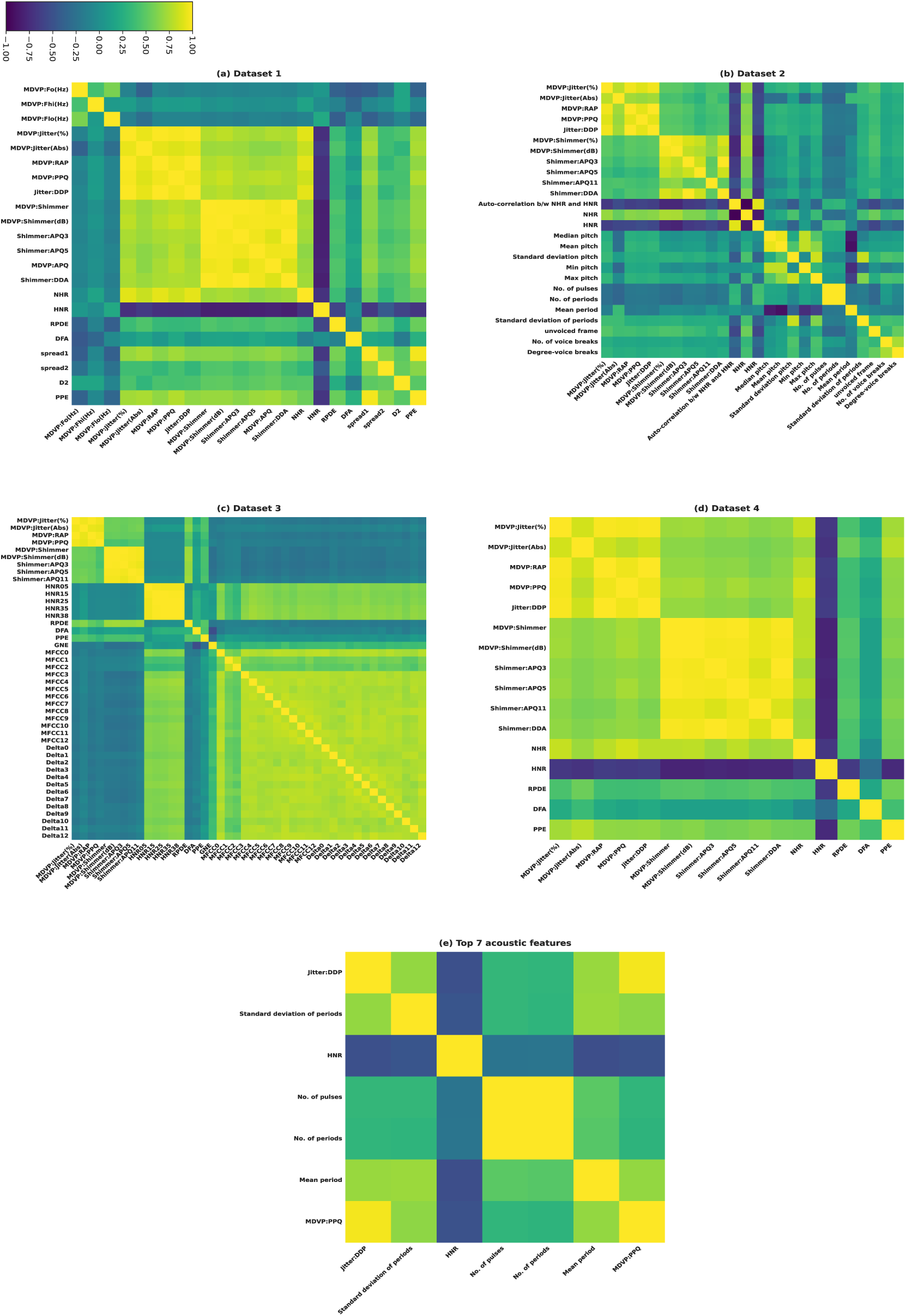
Correlation heatmaps of acoustic features for Datasets 1–4 (a–d), illustrating high redundancy, and the reduced RF–SHAP-selected top feature subset (e), showing minimized multicollinearity.

From the figure, each dataset (a-d) exhibits distinct clusters of highly correlated acoustic features, most notably among jitter measures, shimmer measures, and MFCC coefficients, indicating redundancy within these feature groups. Because several features show strong pairwise correlations across all four datasets, dimensionality reduction is therefore warranted.

**Figure (e)** presents the correlation heatmap of the top seven acoustic features selected using the combined RF–SHAP approach to prevent overfitting associated with the use of all 70 features. The selected features include Jitter:DDP, standard deviation of periods, HNR, number of pulses, number of periods, mean period, and MDVP:PPQ. Most of these features exhibit moderate to low correlation, indicating minimal redundancy, while the number of pulses and number of periods show relatively higher correlation due to their inherent relationship. In contrast, HNR demonstrates weak correlation with other features, suggesting that it captures complementary information relevant to PD detection. Overall, the reduced multicollinearity compared to the original 70-feature set confirms that the selected features are both discriminative and diverse, thereby reducing overfitting risk and improving model generalization. To further evaluate the effectiveness of the selected features, experiments were conducted using 23 machine learning models under two settings: the complete set of 70 acoustic features and the reduced set of 7 features obtained via the RF–SHAP approach. All datasets were combined to form a unified dataset of 7350 samples. Model performance was assessed using four evaluation metrics, namely accuracy, precision, recall, and F1-score, and validated using 10-fold cross-validation. The comparative results are presented in the tables below.

**Table 3.** Classification Performance Using the Entire Acoustic Feature Set (70 Features)

| Model | Accuracy | Precision | Recall | F1-Score |
| --- | --- | --- | --- | --- |
| Logistic Regression | 0.9156 | 0.9988 | 0.9079 | 0.9511 |
| Random Forest | 0.9473 | 0.9654 | 0.9768 | 0.9710 |
| Decision Tree | 0.9400 | 0.9926 | 0.9695 | 0.9800 |
| Support Vector Machine (SVM) | 0.9136 | 0.9988 | 0.9057 | 0.9499 |
| HistGradientBoosting | 0.9481 | 0.9722 | 0.9705 | 0.9713 |
| XGBoost Classifier | 0.9401 | 0.9867 | 0.9466 | 0.9662 |
| LightGBM | 0.9440 | 0.9835 | 0.9542 | 0.9686 |
| CatBoost | 0.9333 | 0.9926 | 0.9334 | 0.9620 |
| Extra Trees | 0.9491 | 0.9681 | 0.9760 | 0.9720 |
| Gradient Boosting | 0.9442 | 0.9645 | 0.9743 | 0.9693 |
| Naïve Bayes | 0.9081 | 0.9878 | 0.9099 | 0.9472 |
| MLP Classifier | 0.9435 | 0.9700 | 0.9675 | 0.9687 |
| Bagging | 0.9474 | 0.9710 | 0.9709 | 0.9709 |
| AdaBoost | 0.9314 | 0.9600 | 0.9709 | 0.9622 |
| Gradient Boosting (Tuned) | 0.9507 | 0.9704 | 0.9754 | 0.9729 |
| LSTM | 0.9380 | 0.9811 | 0.9709 | 0.9659 |
| Voting Ensemble (xgb+lgbm+gboost → LR) | 0.9456 | 0.9830 | 0.9565 | 0.9695 |
| Stacking Ensemble (xgb+lgbm+cat → LR) | 0.9456 | 0.9736 | 0.9662 | 0.9699 |
| Stacking Ensemble (lgbm+cat+mlp → LR) | 0.9340 | 0.9940 | 0.9329 | 0.9624 |
| Voting Ensemble (xgb+rf+hgb → LR) | 0.9463 | 0.9772 | 0.9633 | 0.9702 |
| Stacking Ensemble (xgb+lgbm+cat → LR) | 0.9456 | 0.9736 | 0.9662 | 0.9699 |
| Voting Ensemble (xgb+lgbm+cat → RF) | 0.9429 | 0.9926 | 0.9464 | 0.9678 |
| Stacked (Bagging + AdaBoost) | 0.9469 | 0.9688 | 0.9728 | 0.9708 |

**Tables 3 and 4** present the classification performance of all models using the full feature set and the reduced feature subset, respectively. Using all 70 features (Table 3), the highest accuracy is achieved by the tuned Gradient Boosting model (95.07%), followed by Extra Trees (94.91%), Bagging (94.74%), and Random Forest (94.73%). These models also exhibit strong precision and recall; for example, Random Forest achieves 96.54% precision and 97.68% recall (F1-score: 97.10%), while Extra Trees attains an F1-score of 97.20%. However, this high performance can be attributed to feature redundancy and multicollinearity, as observed in the correlation analysis.

**Table 4.** Performance Evaluation Using the Top 7 Acoustic Features Selected via RF–SHAP.

| Model | Accuracy | Precision | Recall |
| --- | --- | --- | --- |
| Logistic Regression | 0.8943 | 0.9915 | 0.8911 |
| Random Forest | 0.9290 | 0.9551 | 0.9671 |
| Decision Tree | 0.9165 | 0.9500 | 0.9553 |
| Support Vector Machine (SVM) | 0.8933 | 0.9915 | 0.8900 |
| HistGradientBoosting | 0.9248 | 0.9548 | 0.9626 |
| XGBoost Classifier | 0.9063 | 0.9900 | 0.9057 |
| LightGBM | 0.9163 | 0.9820 | 0.9245 |
| CatBoost | 0.9149 | 0.9878 | 0.9174 |
| Extra Trees | 0.9287 | 0.9573 | 0.9643 |
| Gradient Boosting | 0.9280 | 0.9586 | 0.9621 |
| Naïve Bayes | 0.9069 | 0.9694 | 0.9256 |
| MLP Classifier | 0.9241 | 0.9539 | 0.9628 |
| Bagging | 0.9261 | 0.9577 | 0.9609 |
| AdaBoost | 0.9139 | 0.9467 | 0.9595 |
| Gradient Boosting (Tuned) | 0.9283 | 0.9569 | 0.9629 |
| LSTM | 0.9236 | 0.9535 | 0.9630 |
| Voting Ensemble (xgb+lgbm+gboost → LR) | 0.9194 | 0.9795 | 0.9306 |
| Stacking Ensemble (xgb+lgbm+cat → LR) | 0.9252 | 0.9654 | 0.9516 |
| Stacking Ensemble (lgbm+cat+mlp → LR) | 0.9133 | 0.9903 | 0.9133 |
| Voting Ensemble (xgb+rf+hgb → LR) | 0.9257 | 0.9685 | 0.9490 |
| Stacking Ensemble (xgb+lgbm+cat → LR) | 0.9252 | 0.9654 | 0.9516 |
| Voting Ensemble (xgb+lgbm+cat → RF) | 0.9190 | 0.9853 | 0.9246 |
| Stacked (Bagging + AdaBoost) | 0.9211 | 0.9493 | 0.9646 |

In contrast, Table 4 highlights that the top 7 features selected using the RF–SHAP approach achieve highly competitive performance despite significant dimensionality reduction, with Random Forest (92.90%), Extra Trees (92.87%), and Gradient Boosting (92.80%) maintaining strong F1-scores of 96.10%, 96.08%, and 96.03%, respectively. Notably, Logistic Regression achieves 89.43% accuracy with very high precision (99.15%) and an F1-score of 93.85%, indicating stable and reliable predictions. Across all models, precision, recall, and F1-scores remain consistently high and well-balanced, demonstrating that the reduced feature set effectively preserves the essential discriminatory information. Overall, the compact feature set delivers near-equivalent predictive performance, highlighting the effectiveness of the RF–SHAP approach in reducing redundancy while improving generalization, interpretability, and computational efficiency without meaningful performance degradation.

### 3.2 MEG Feature Analysis, Selection, and Classification Performance

Following the acoustic analysis, a similar procedure is applied to the MEG data across all three tasks. Correlation analysis is first performed on the full set of 21 MEG features (7350 × 21) to assess redundancy and inter-feature dependencies. Subsequently, RF–SHAP-based feature selection is employed to obtain a reduced subset of 7 discriminative features (7350 × 7). The resulting correlation patterns for both the original and reduced feature sets are illustrated below.

**Figure S2.**
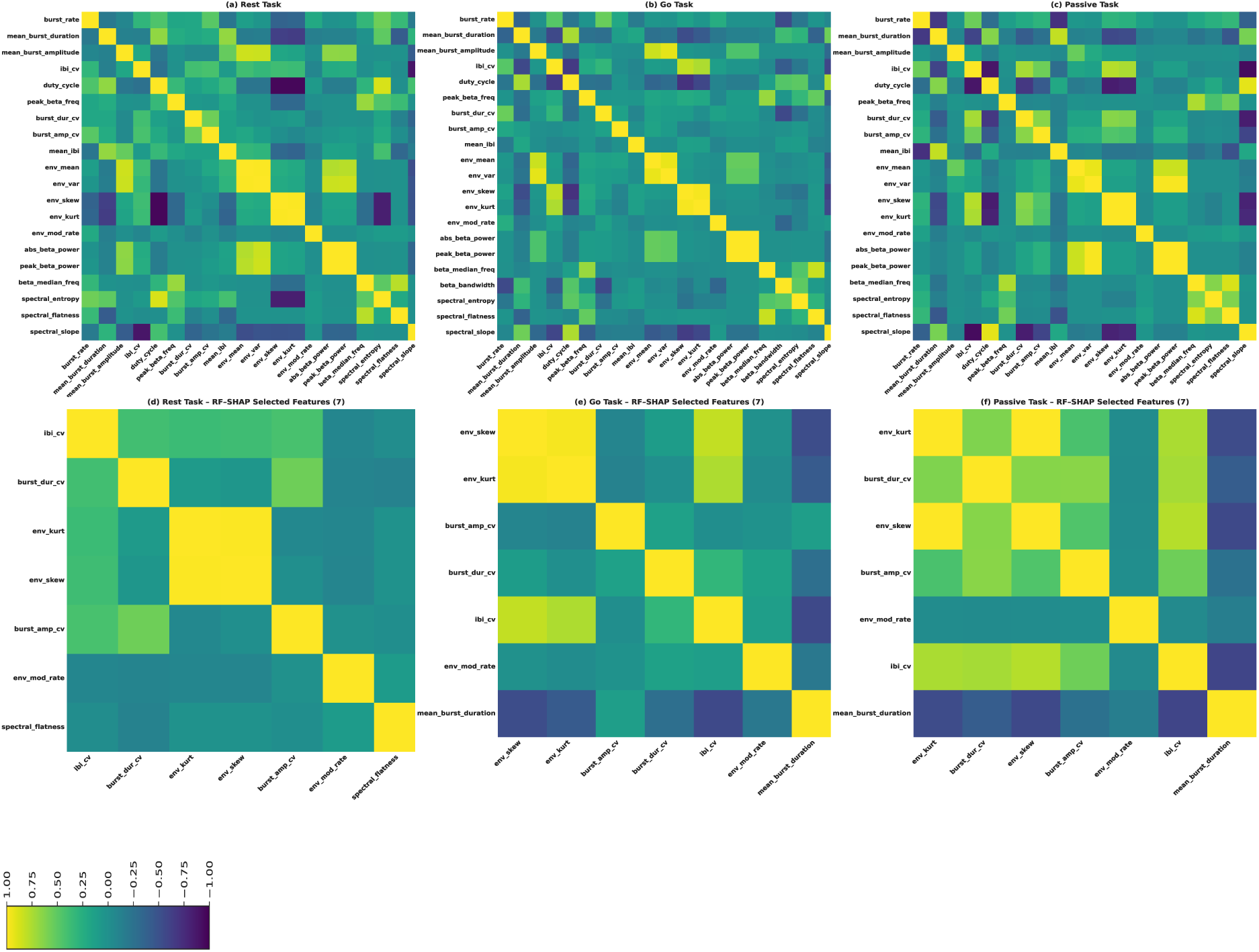
Correlation heatmaps of MEG features across Rest, Go, and Passive tasks. (a–c) depict the full feature set after removal of zero-variance features. (d–f) present the reduced feature subsets obtained using the RF–SHAP approach, demonstrating decreased redundancy and improved feature independence across all conditions, with the Go task showing a more balanced correlation structure.

The correlation heatmaps presented in Fig. S2 illustrate the inter-feature relationships of MEG signals across three experimental conditions: Rest, Go, and Passive tasks. The top row (a–c) represents the full feature set, where features exhibiting zero variance were removed prior to analysis to avoid undefined correlations. From the figure, it is evident that all three tasks exhibit notable clusters of moderate to high correlations, particularly among statistical features such as the mean, variance, skewness, and kurtosis of the envelope, as well as burst-related features. Among the three conditions, the Go task (b) demonstrates a well-structured correlation pattern, capturing strong yet organized inter-feature relationships, which may contribute to improved discriminative capability. In contrast, the Passive task (c) exhibits more widespread correlations, indicating higher redundancy among features, whereas the Rest task (a) shows comparatively moderate inter-feature dependencies.

The bottom row (d–f) presents the correlation structure of the reduced feature subsets obtained using the RF–SHAP method. Although the top seven features differ slightly across tasks, several features consistently appear in all three conditions, including env_skew, env_kurt, burst_amp_cv, burst_dur_cv, ibi_cv, and env_mod_rate, highlighting their robustness and relevance for Parkinson’s disease discrimination. Task-specific variations are also observed; for example, the Rest task additionally includes spectral_flatness, whereas the Go and Passive tasks include mean_burst_duration, indicating subtle differences in feature importance across cognitive states. Furthermore, the reduced feature sets exhibit significantly lower inter-feature correlations compared to the full feature sets, demonstrating effective mitigation of multicollinearity. Among the three conditions, the Go task (e) shows a more balanced and less redundant correlation structure, while the Passive task (f) exhibits relatively higher correlation between env_skew and env_kurt, reflecting their inherent statistical dependence. In contrast, the Rest task (d) shows minimal strong correlations among the selected features.

Overall, the RF–SHAP-based feature selection effectively reduces feature redundancy while preserving the most discriminative and complementary features. This not only improves interpretability but also enhances the robustness and generalization capability of downstream classification models.

To comprehensively evaluate the discriminative capability of MEG-derived biomarkers, two complementary representation strategies were investigated: (i) channel-level MEG representations, which preserve sensor-specific neural information, and (ii) participant-level hemispheric aggregation representations, which summarize neural activity at the hemispheric level. The corresponding analyses and classification results for both representations are presented in the following sections.

#### i) Channel- wise MEG analysis

Using the unified MEG dataset consisting of 7350 samples, model performance was evaluated on both the complete feature set (7350 × 21) and the reduced feature set (7350 x 7) derived via the RF–SHAP method across all three task conditions. The corresponding comparative results are illustrated in the tables below.

**Table (a).** Classification Performance Under Rest Condition — Full Feature Set (7350 × 21), 10-fold CV, 80:20 split.

| Model | Accuracy | Precision | Recall | F1-Score |
| --- | --- | --- | --- | --- |
| Logistic Regression | 0.6361 | 0.6414 | 0.5803 | 0.6092 |
| Random Forest | 0.7919 | 0.8183 | 0.7392 | 0.7764 |
| Decision Tree | 0.7321 | 0.7324 | 0.7127 | 0.7222 |
| Support Vector Machine (SVM) | 0.6397 | 0.6680 | 0.5231 | 0.5865 |
| HistGradientBoosting | 0.7957 | 0.8133 | 0.7558 | 0.7833 |
| XGBoost Classifier | 0.7979 | 0.8037 | 0.7764 | 0.7895 |
| LightGBM | 0.7918 | 0.8011 | 0.7656 | 0.7822 |
| CatBoost | 0.7971 | 0.8092 | 0.7675 | 0.7872 |
| Extra Trees | 0.7763 | 0.7992 | 0.7266 | 0.7606 |
| Gradient Boosting | 0.7484 | 0.7658 | 0.7021 | 0.7319 |
| Naïve Bayes | 0.4977 | 0.4935 | 0.9910 | 0.6586 |
| MLP Classifier | 0.7195 | 0.7333 | 0.6791 | 0.7024 |
| Bagging | 0.7982 | 0.8145 | 0.7619 | 0.7869 |
| AdaBoost | 0.6945 | 0.7059 | 0.6451 | 0.6738 |
| Gradient Boosting (Tuned) | 0.7758 | 0.7920 | 0.7350 | 0.7622 |
| LSTM | 0.6780 | 0.6936 | 0.6194 | 0.6527 |
| Voting Ensemble (xgb+lgbm+gboost<br>→ LR) | 0.7946 | 0.8091 | 0.7611 | 0.7839 |
| Stacking Ensemble (xgb+lgbm+cat<br>→ LR) | 0.7963 | 0.8079 | 0.7658 | 0.7861 |
| Stacking Ensemble (lgbm+cat+mlp<br>→ LR) | 0.7906 | 0.7952 | 0.7710 | 0.7826 |
| Voting Ensemble (xgb+rf+hgb → LR) | 0.7966 | 0.8172 | 0.7529 | 0.7834 |
| Stacking Ensemble (xgb+lgbm+cat → LR) | 0.7963 | 0.8079 | 0.7658 | 0.7861 |
| Voting Ensemble (xgb+lgbm+cat → RF) | 0.7937 | 0.8089 | 0.7579 | 0.7823 |
| Stacked (Bagging + AdaBoost) | 0.7917 | 0.8031 | 0.7613 | 0.7813 |

**Table (b).** Classification Performance Under Go Condition — Full Feature Set (7350 × 21), 10-fold CV, 80:20 split.

| Model | Accuracy | Precision | Recall | F1-Score |
| --- | --- | --- | --- | --- |
| Logistic Regression | 0.5801 | 0.5620 | 0.6401 | 0.5984 |
| Random Forest | 0.6793 | 0.6790 | 0.6537 | 0.6658 |
| Decision Tree | 0.6134 | 0.6069 | 0.5962 | 0.6014 |
| Support Vector Machine (SVM) | 0.5715 | 0.5492 | 0.6923 | 0.6124 |
| HistGradientBoosting | 0.6848 | 0.6751 | 0.6859 | 0.6802 |
| XGBoost Classifier | 0.6734 | 0.6635 | 0.6750 | 0.6690 |
| LightGBM | 0.6685 | 0.6544 | 0.6848 | 0.6687 |
| CatBoost | 0.6656 | 0.6518 | 0.6808 | 0.6658 |
| Extra Trees | 0.6683 | 0.6617 | 0.6589 | 0.6599 |
| Gradient Boosting | 0.6384 | 0.6238 | 0.6571 | 0.6398 |
| Naïve Bayes | 0.5073 | 0.4983 | 0.9978 | 0.6644 |
| MLP Classifier | 0.6554 | 0.6461 | 0.6532 | 0.6482 |
| Bagging | 0.6613 | 0.6568 | 0.6445 | 0.6503 |
| AdaBoost | 0.5731 | 0.5729 | 0.5124 | 0.5343 |
| Gradient Boosting (Tuned) | 0.6634 | 0.6545 | 0.6608 | 0.6572 |
| LSTM | 0.6256 | 0.6135 | 0.6320 | 0.6220 |
| Voting Ensemble (xgb+lgbm+gboost<br>→ LR) | 0.6699 | 0.6572 | 0.6798 | 0.6681 |
| Stacking Ensemble (xgb+lgbm+cat<br>→ LR) | 0.6776 | 0.6711 | 0.6702 | 0.6703 |
| Stacking Ensemble (lgbm+cat+mlp<br>→ LR) | 0.6813 | 0.6731 | 0.6776 | 0.6750 |
| Voting Ensemble (xgb+rf+hgb →<br>LR) | 0.6733 | 0.6671 | 0.6642 | 0.6654 |
| Stacking Ensemble (xgb+lgbm+cat<br>→ LR) | 0.6776 | 0.6711 | 0.6702 | 0.6703 |
| Voting Ensemble (xgb+lgbm+cat →<br>RF) | 0.6752 | 0.6655 | 0.6768 | 0.6708 |
| Stacked Ensemble (Bagging +<br>AdaBoost) | 0.6689 | 0.6615 | 0.6640 | 0.6623 |

**Table (c).**
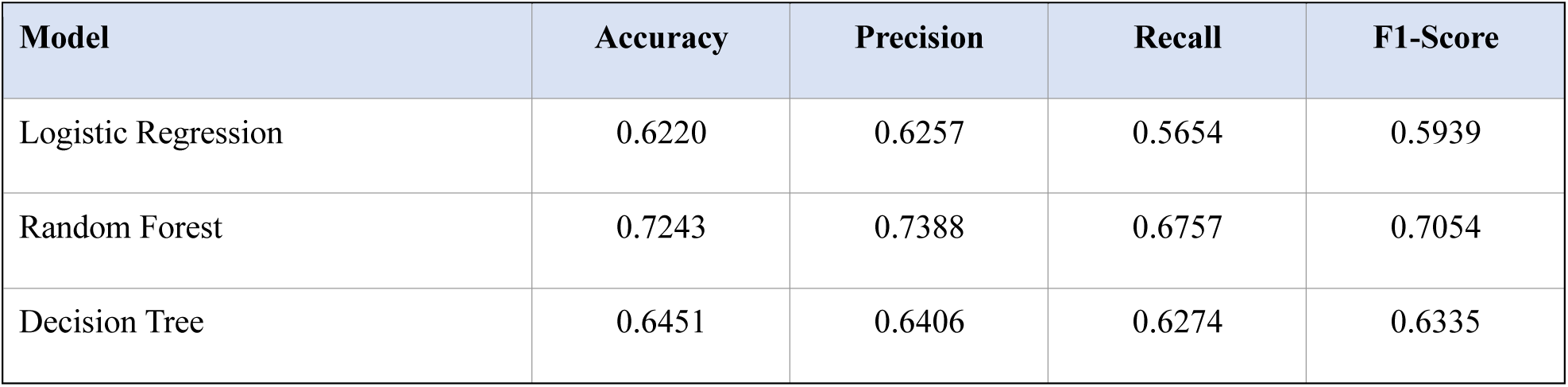
Classification Performance Under Passive Task Condition — Full Feature Set (7350 × 21), 10-fold CV, 80:20 split.

**Table 5.**
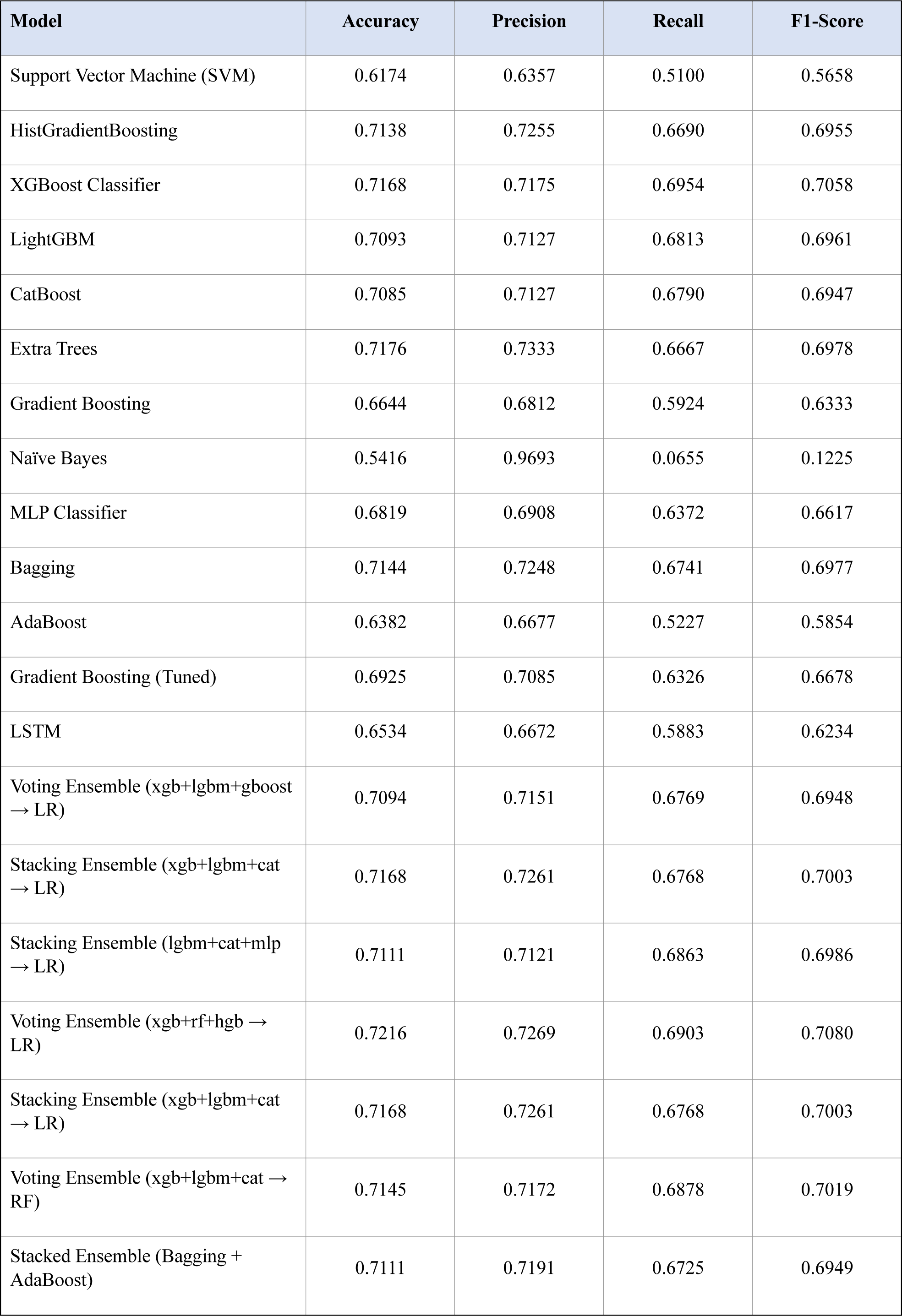
Classification performance of machine learning models using the full feature set (7350 × 21) under (a) Rest, (b) Go, and (c) Passive task conditions, evaluated using 10-fold cross-validation with an 80:20 split in terms of accuracy, precision, recall, and F1-score.

**Tables (a)–(c)** present the classification performance of 23 machine learning models evaluated using the full MEG feature set under (a) Rest, (b) Go, and (c) Passive task conditions. These results are consistent with the correlation patterns observed in Fig. S2, where the Go task demonstrates superior performance. Specifically, the Go condition achieves the highest overall accuracy of 0.7982 with the Bagging classifier, closely followed by XGBoost (0.7979) and several ensemble models exceeding 0.79. In terms of other evaluation metrics, the Go task also shows strong performance, with precision reaching up to 0.8183 (Random Forest), recall up to 0.7764 (XGBoost), and F1-score up to 0.7895 (XGBoost), indicating enhanced discriminative capability. In comparison, the Passive task exhibits moderate performance, with the highest accuracy of 0.7243 achieved by Random Forest, followed by competitive results from ensemble methods such as Voting (0.7216) and XGBoost (0.7168). The corresponding precision (up to 0.7388), recall (up to 0.6954), and F1-score (up to 0.7080) indicate stable but comparatively lower performance than the Go task. In contrast, the Rest task shows the lowest classification performance, with a maximum accuracy of 0.6848 obtained using HistGradientBoosting, while most other models achieve accuracies below 0.68. Similarly, other evaluation metrics remain relatively lower; although recall is occasionally high (e.g., Naïve Bayes), it is accompanied by poor precision, resulting in imbalanced F1-scores. Overall, these results indicate that the Go task provides the most informative and discriminative MEG patterns for Parkinson’s disease classification, followed by the Passive and Rest conditions. The subsequent table illustrates the classification performance using the reduced feature subset (7350 × 7) obtained through the RF–SHAP approach across all three task conditions.

**Table (a).** Classification Performance Under Rest Condition — Reduced Feature Set (7350 × 7) via RF–SHAP.

| Model | Accuracy | Precision | Recall | F1-Score |
| --- | --- | --- | --- | --- |
| Logistic Regression | 0.5820 | 0.5719 | 0.5764 | 0.5740 |
| Random Forest | 0.6512 | 0.6460 | 0.6354 | 0.6405 |
| Decision Tree | 0.6000 | 0.5949 | 0.5711 | 0.5826 |
| Support Vector Machine (SVM) | 0.5804 | 0.5795 | 0.5155 | 0.5455 |
| HistGradientBoosting | 0.6419 | 0.6309 | 0.6455 | 0.6379 |
| XGBoost Classifier | 0.6312 | 0.6206 | 0.6315 | 0.6260 |
| LightGBM | 0.6292 | 0.6141 | 0.6530 | 0.6326 |
| CatBoost | 0.6341 | 0.6164 | 0.6685 | 0.6410 |
| Extra Trees | 0.6241 | 0.6159 | 0.6176 | 0.6162 |
| Gradient Boosting | 0.6211 | 0.6064 | 0.6440 | 0.6242 |
| Naïve Bayes | 0.5130 | 0.5013 | 0.9720 | 0.6612 |
| MLP Classifier | 0.6335 | 0.6231 | 0.6424 | 0.6314 |
| Bagging | 0.6261 | 0.6198 | 0.6112 | 0.6150 |
| AdaBoost | 0.5605 | 0.5562 | 0.5399 | 0.5434 |
| Gradient Boosting (Tuned) | 0.6388 | 0.6284 | 0.6374 | 0.6328 |
| LSTM | 0.6151 | 0.5998 | 0.6438 | 0.6204 |
| Voting Ensemble (xgb+lgbm+gboost<br>→ LR) | 0.6332 | 0.6183 | 0.6543 | 0.6353 |
| Stacking Ensemble (xgb+lgbm+cat<br>→ LR) | 0.6359 | 0.6242 | 0.6433 | 0.6334 |
| Stacking Ensemble (lgbm+cat+mlp<br>→ LR) | 0.6429 | 0.6260 | 0.6736 | 0.6482 |
| Voting Ensemble (xgb+rf+hgb →<br>LR) | 0.6340 | 0.6246 | 0.6314 | 0.6278 |
| Stacking Ensemble (xgb+lgbm+cat<br>→ LR) | 0.6359 | 0.6242 | 0.6433 | 0.6334 |
| Voting Ensemble (xgb+lgbm+cat →<br>RF) | 0.6311 | 0.6181 | 0.6440 | 0.6306 |
| Stacked Ensemble (Bagging +<br>AdaBoost) | 0.6367 | 0.6265 | 0.6384 | 0.6322 |

**Table (b).** Classification Performance Under Go task Condition — Reduced Feature Set (7350 × 7) via RF–SHAP.

| Model | Accuracy | Precision | Recall | F1-Score |
| --- | --- | --- | --- | --- |
| Logistic Regression | 0.6179 | 0.6555 | 0.4634 | 0.5423 |
| Random Forest | 0.7416 | 0.7633 | 0.6834 | 0.7208 |
| Decision Tree | 0.7099 | 0.7082 | 0.6939 | 0.7004 |
| Support Vector Machine (SVM) | 0.5814 | 0.7277 | 0.2321 | 0.3513 |
| HistGradientBoosting | 0.7410 | 0.7652 | 0.6791 | 0.7194 |
| XGBoost Classifier | 0.7336 | 0.7385 | 0.7050 | 0.7210 |
| LightGBM | 0.7460 | 0.7661 | 0.6935 | 0.7277 |
| CatBoost | 0.7496 | 0.7708 | 0.6961 | 0.7312 |
| Extra Trees | 0.7505 | 0.7730 | 0.6956 | 0.7319 |
| Gradient Boosting | 0.7246 | 0.7479 | 0.6617 | 0.7017 |
| Naïve Bayes | 0.5113 | 0.5200 | 0.0260 | 0.0495 |
| MLP Classifier | 0.6777 | 0.7167 | 0.5722 | 0.6337 |
| Bagging | 0.7465 | 0.7628 | 0.7009 | 0.7301 |
| AdaBoost | 0.6772 | 0.6874 | 0.6278 | 0.6552 |
| Gradient Boosting (Tuned) | 0.7348 | 0.7548 | 0.6785 | 0.7142 |
| LSTM | 0.6455 | 0.6740 | 0.5469 | 0.5984 |
| Voting Ensemble (xgb+lgbm+gboost<br>→ LR) | 0.7473 | 0.7706 | 0.6900 | 0.7277 |
| Stacking Ensemble (xgb+lgbm+cat<br>→ LR) | 0.7437 | 0.7608 | 0.6951 | 0.7259 |
| Stacking Ensemble (lgbm+cat+mlp<br>→ LR) | 0.7434 | 0.7567 | 0.7013 | 0.7274 |
| Voting Ensemble (xgb+rf+hgb →<br>LR) | 0.7432 | 0.7610 | 0.6922 | 0.7248 |
| Stacking Ensemble (xgb+lgbm+cat<br>→ LR) | 0.7437 | 0.7608 | 0.6951 | 0.7259 |
| Voting Ensemble (xgb+lgbm+cat → RF) | 0.7439 | 0.7591 | 0.6982 | 0.7270 |
| Stacked (Bagging + AdaBoost) | 0.7372 | 0.7533 | 0.6880 | 0.7188 |

**Table (c).**
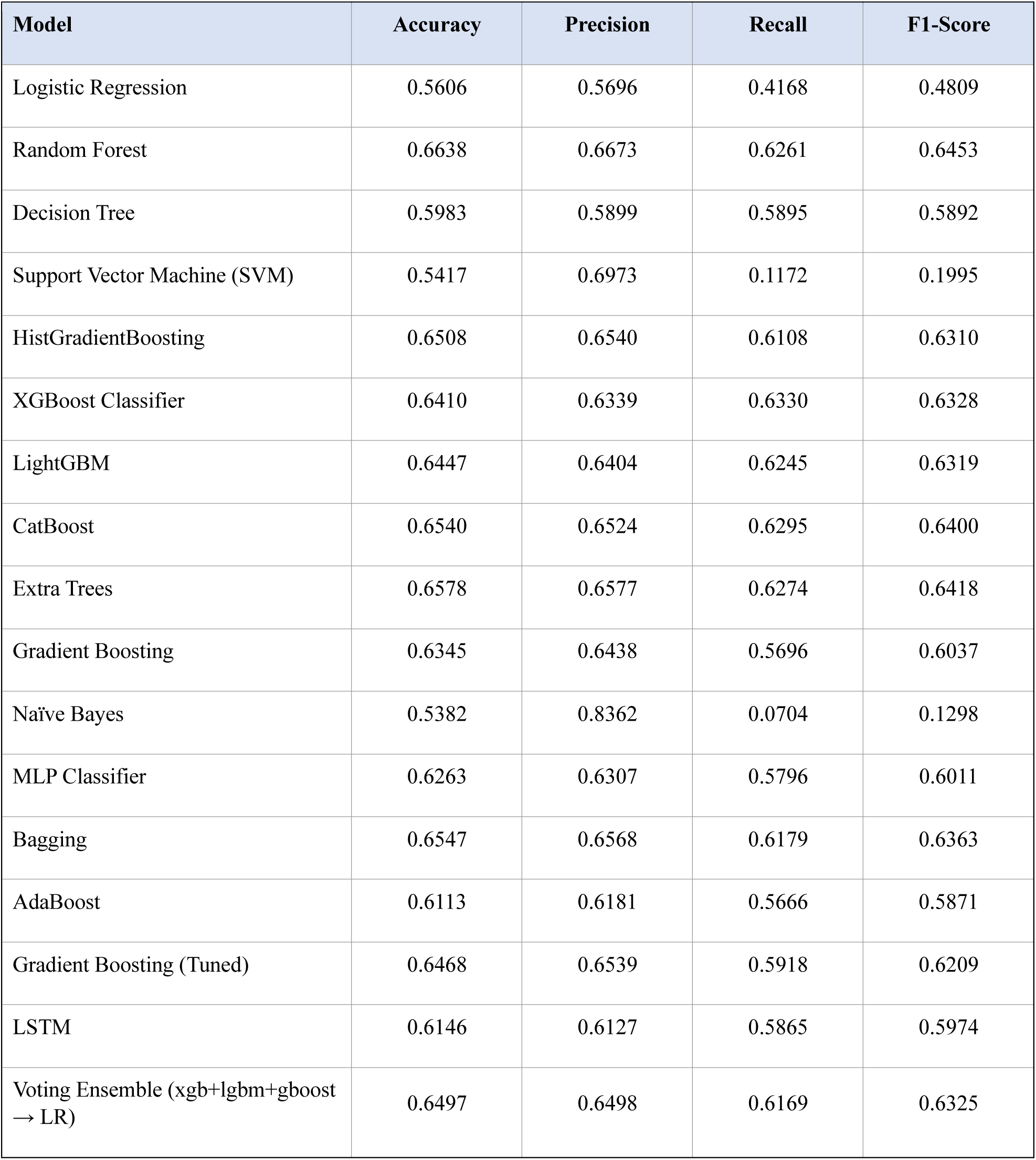
Classification Performance Under Passive Task Condition — Reduced Feature Set (7350 × 7) via RF–SHAP.

**Table 6.**
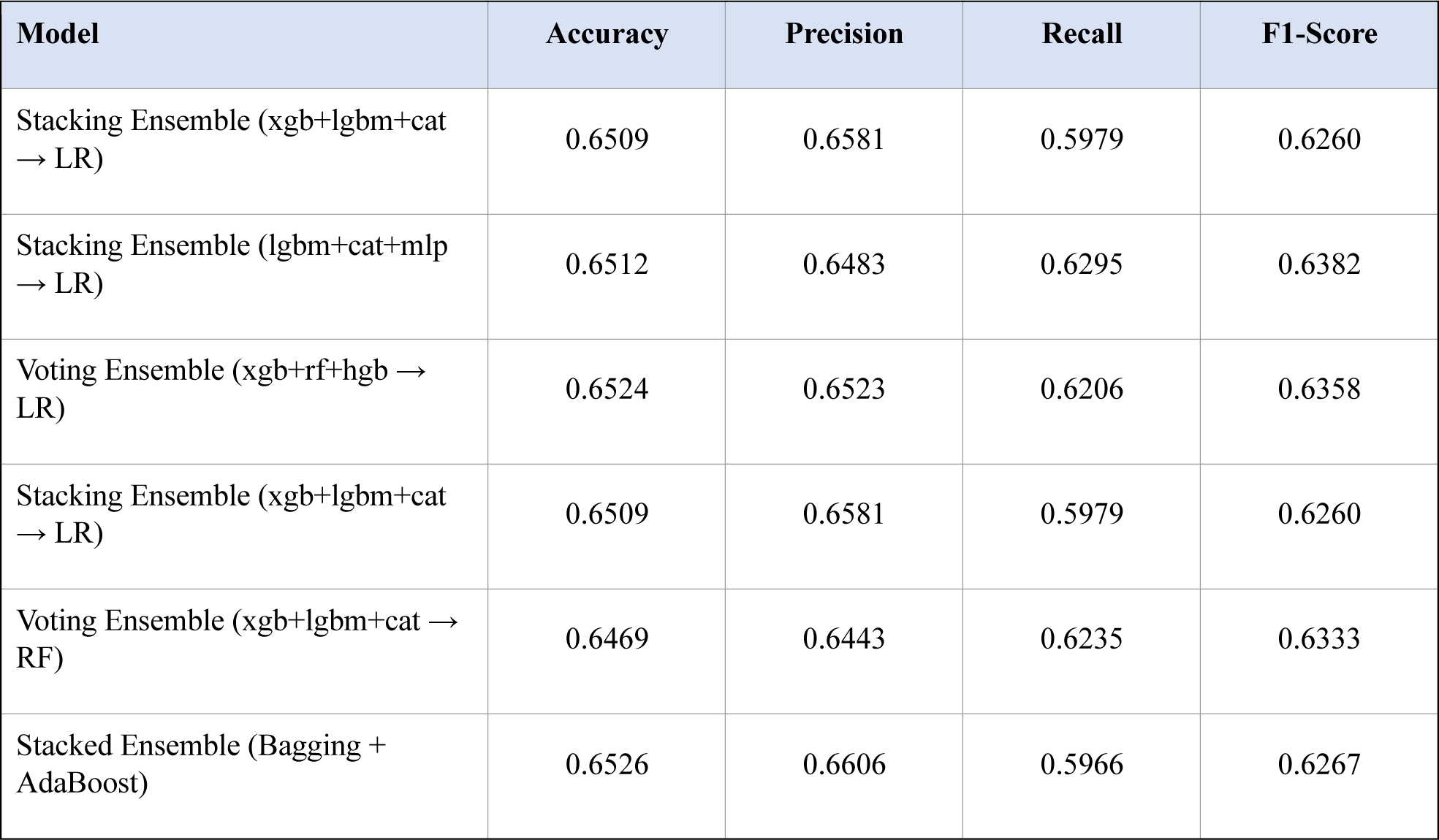
Classification performance of machine learning models using the reduced feature set (7350 × 7) obtained via RF–SHAP across (a) Rest, (b) Go, and (c) Passive task conditions, evaluated using 10-fold cross-validation with an 80:20 split in terms of accuracy, precision, recall, and F1-score.

Tables (a)–(c) present the classification performance of models using the reduced feature subset (7350 × 7) across (a) Rest, (b) Go, and (c) Passive task conditions. These results are consistent with both the full feature set results reported in **Table 5** and the correlation analysis shown in **Fig. S2**, where the Go task continues to demonstrate superior performance. Specifically, the Go condition achieves the highest accuracy of 0.7505 with Extra Trees, closely followed by CatBoost (0.7496), Voting ensembles (∼0.7473), and Bagging (0.7465). In terms of other evaluation metrics, the Go task also outperforms the other conditions, with precision reaching up to 0.7730, recall up to 0.7050 (XGBoost), and F1-score up to 0.7319 (Extra Trees), indicating that the well-structured and less redundant feature relationships observed in the correlation heatmaps translate into effective classification performance. In comparison, the Passive task exhibits moderate performance, with the highest accuracy of 0.6638 (Random Forest), followed by ensemble methods such as Voting (0.6524) and Bagging (0.6547). The corresponding precision (up to 0.6673), recall (up to 0.6330), and F1-score (up to 0.6453) indicate stable but comparatively lower performance than the Go task. In contrast, the Rest task shows the lowest performance, with a maximum accuracy of 0.6512 (Random Forest) and competitive results from stacking-based ensembles (0.6429), while most other models remain at or below this range. When compared to the full feature set results, the reduced RF–SHAP feature subset shows only a marginal decrease in performance across all three tasks, while significantly reducing feature dimensionality and multicollinearity. This demonstrates that the RF–SHAP approach effectively preserves the most discriminative and complementary information, as also supported by the reduced correlation patterns, thereby maintaining robust classification performance with a more compact and interpretable feature space.

#### ii) Participant-Level Analysis Using Hemispheric Aggregation

To complement the channel-level analysis, a participant-level MEG representation was constructed through hemispheric aggregation. The 306 MEG sensors were grouped into left and right hemispheric regions according to their spatial positions in the Elekta Neuromag sensor layout. For each feature, hemisphere-specific means, standard deviations, and normalized asymmetry indices were computed, yielding participant-level descriptors that summarize hemispheric neural dynamics and interhemispheric differences. The classification results obtained using these hemispheric aggregation features are presented in the following tables.

**Table 7a.** Classification Performance Using Participant-Level Hemispheric Aggregation Features Under Rest Condition (80:20 split, 10-fold CV)

| Model | Accuracy | Precision | Recall | F1-Score |
| --- | --- | --- | --- | --- |
| Logistic Regression | 0.5525 | 0.5586 | 0.5893 | 0.5354 |
| Random Forest | 0.5769 | 0.7500 | 0.5294 | 0.6206 |
| Decision Tree | 0.4230 | 0.5833 | 0.4117 | 0.4827 |
| Support Vector Machine (SVM) | 0.6000 | 0.6046 | 0.6854 | 0.6046 |
| HistGradientBoosting | 0.6467 | 0.6354 | 0.6529 | 0.6315 |
| XGBoost Classifier | 0.5762 | 0.5680 | 0.6321 | 0.5772 |
| LightGBM | 0.5945 | 0.5250 | 0.4714 | 0.4820 |
| CatBoost | 0.5945 | 0.4883 | 0.5138 | 0.4901 |
| Extra Trees | 0.6045 | 0.5050 | 0.4221 | 0.4502 |
| Gradient Boosting | 0.5845 | 0.5107 | 0.4340 | 0.4549 |
| Naïve Bayes | 0.4763 | 0.4615 | 0.9800 | 0.6144 |
| MLP Classifier | 0.5145 | 0.4380 | 0.4464 | 0.4287 |
| Bagging | 0.5654 | 0.4391 | 0.3833 | 0.3966 |
| AdaBoost | 0.5754 | 0.5471 | 0.4554 | 0.4577 |
| Gradient Boosting (Tuned) | 0.6154 | 0.8182 | 0.5294 | 0.6429 |
| LSTM | 0.5745 | 0.4616 | 0.4873 | 0.4669 |
| Voting Ensemble (xgb+lgbm+gboost<br>→ LR) | 0.5945 | 0.5517 | 0.5114 | 0.5014 |
| Stacking Ensemble (xgb+lgbm+cat → LR) | 0.5064 | 0.5132 | 0.5592 | 0.4872 |
| Stacking Ensemble (lgbm+cat+mlp → LR) | 0.5536 | 0.5083 | 0.5583 | 0.5164 |
| Voting Ensemble (xgb+rf+hgb → LR) | 0.5227 | 0.5269 | 0.5333 | 0.4984 |
| Stacking Ensemble (xgb+lgbm+cat → LR) | 0.5064 | 0.5132 | 0.5592 | 0.4872 |
| Voting Ensemble (xgb+lgbm+cat → RF) | 0.5027 | 0.5019 | 0.5208 | 0.4817 |
| Stacked Ensemble (Bagging + AdaBoost) | 0.4973 | 0.5008 | 0.5442 | 0.4740 |

**Table 7b.** Classification Performance Using Participant-Level Hemispheric Aggregation Features Under Go Condition (80:20 split, 10-fold CV)

| Model | Accuracy | Precision | Recall | F1-Score |
| --- | --- | --- | --- | --- |
| Logistic Regression | 0.6487 | 0.6461 | 0.6104 | 0.6064 |
| Random Forest | 0.7096 | 0.7304 | 0.6802 | 0.6716 |
| Decision Tree | 0.5916 | 0.5361 | 0.5214 | 0.5120 |
| Support Vector Machine (SVM) | 0.6660 | 0.6633 | 0.5915 | 0.6103 |
| HistGradientBoosting | 0.7487 | 0.7139 | 0.6802 | 0.6838 |
| XGBoost Classifier | 0.6807 | 0.6510 | 0.6292 | 0.6127 |
| LightGBM | 0.6709 | 0.6966 | 0.6254 | 0.6243 |
| CatBoost | 0.5909 | 0.5900 | 0.5138 | 0.5060 |
| Extra Trees | 0.6400 | 0.6371 | 0.5478 | 0.5618 |
| Gradient Boosting | 0.6309 | 0.6661 | 0.5730 | 0.5687 |
| Naïve Bayes | 0.4854 | 0.4177 | 0.8550 | 0.5522 |
| MLP Classifier | 0.6118 | 0.5564 | 0.5847 | 0.5476 |
| Bagging | 0.6318 | 0.6083 | 0.5697 | 0.5591 |
| AdaBoost | 0.6618 | 0.6700 | 0.6061 | 0.6017 |
| Gradient Boosting (Tuned) | 0.7583 | 0.7560 | 0.7253 | 0.7195 |
| LSTM | 0.6509 | 0.6421 | 0.5321 | 0.5401 |
| Voting Ensemble (xgb+lgbm+gboost → LR) | 0.6909 | 0.7300 | 0.6431 | 0.6519 |
| Stacking Ensemble (xgb+lgbm+cat → LR) | 0.6355 | 0.6567 | 0.5958 | 0.5949 |
| Stacking Ensemble (lgbm+cat+mlp → LR) | 0.6418 | 0.6083 | 0.5533 | 0.5652 |
| Voting Ensemble (xgb+rf+hgb → LR) | 0.7018 | 0.7583 | 0.5683 | 0.6417 |
| Stacking Ensemble (xgb+lgbm+cat → LR) | 0.6355 | 0.6567 | 0.5958 | 0.5949 |
| Voting Ensemble (xgb+lgbm+cat → RF) | 0.7218 | 0.7798 | 0.6133 | 0.6738 |
| Stacked (Bagging + AdaBoost) | 0.6655 | 0.6821 | 0.6333 | 0.6297 |

**Table 7c.**
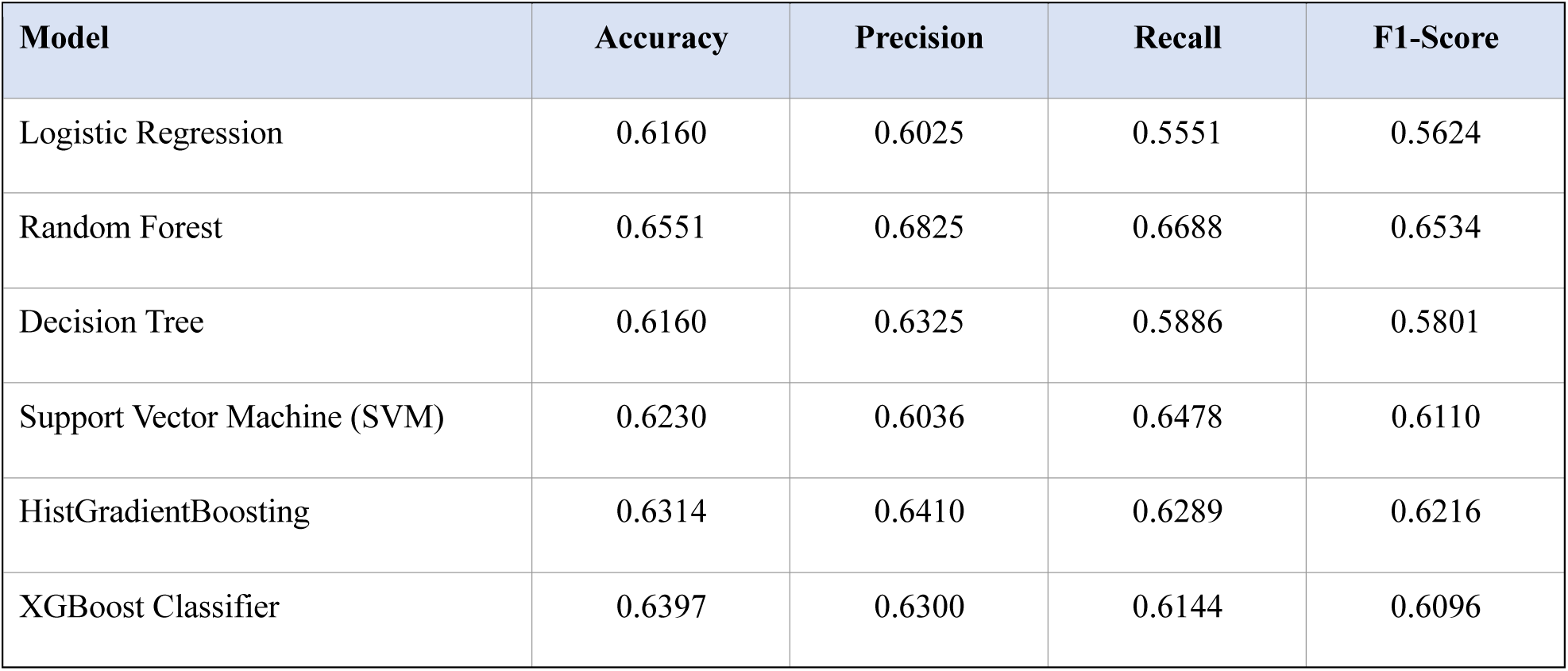
Classification Performance Using Participant-Level Hemispheric Aggregation Features Under Passive Task Condition (80:20 split, 10-fold CV)

**Table 7.**
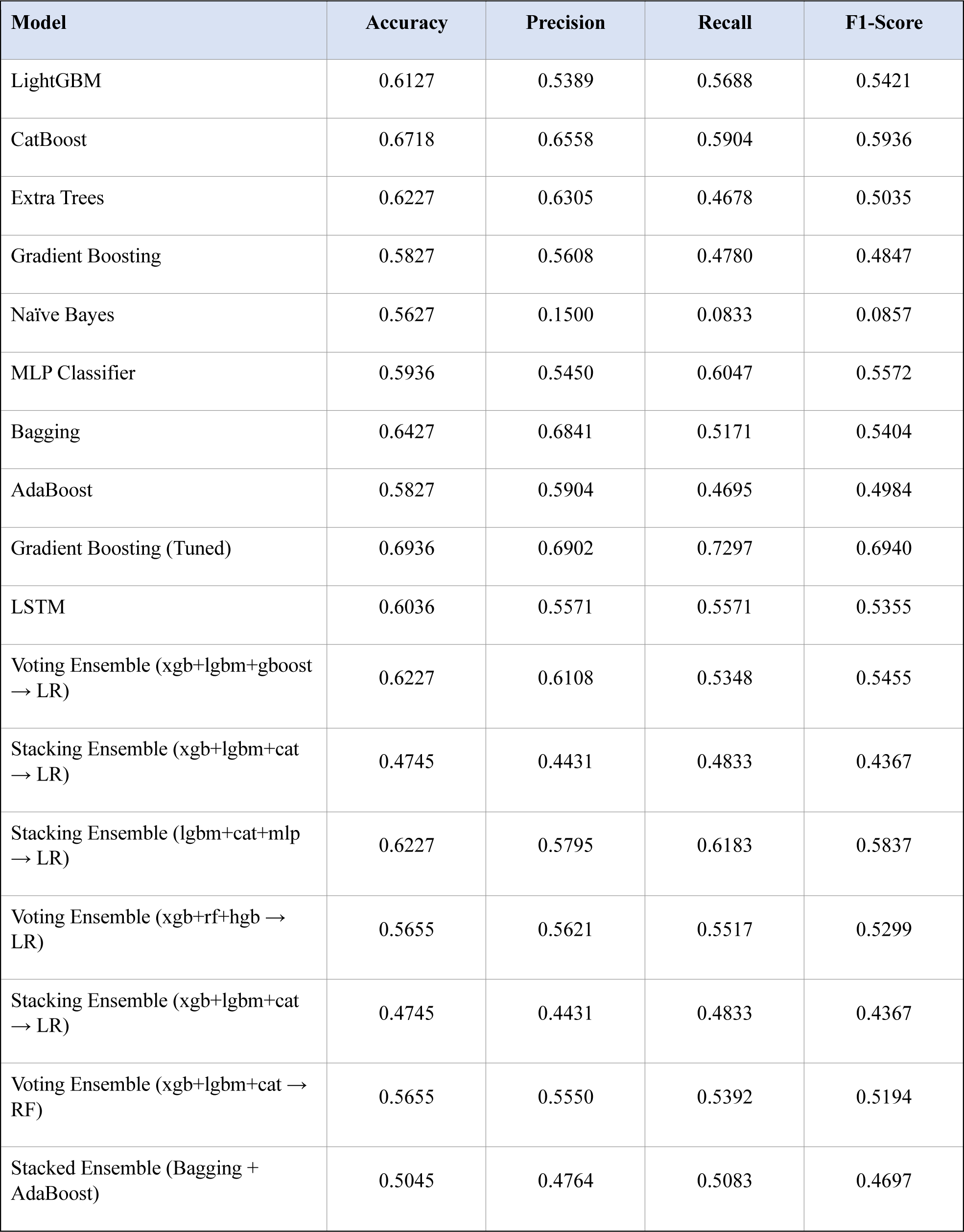
Classification performance of machine learning models using participant-level hemispheric aggregation features across (a) Rest, (b) Go, and (c) Passive task conditions, evaluated using an 80:20 train-test split with 10-fold cross-validation. Performance is reported in terms of Accuracy, Precision, Recall, and F1-score.

As shown in **Table 7**, participant-level hemispheric aggregation yielded moderate classification performance across all three task conditions. Among the evaluated paradigms, the Go task demonstrated the strongest discriminative capability, achieving a maximum accuracy of 75.83% and an F1-score of 71.95% using the tuned Gradient Boosting classifier. The Passive task exhibited intermediate performance, with a peak accuracy of 69.36% and F1-score of 69.40%, whereas the Rest task produced comparatively lower results, reaching a maximum accuracy of 64.67% and F1-score of 64.29%. However, when compared with the channel-level analyses reported in **Tables 5** and **6**, hemispheric aggregation consistently resulted in lower classification performance. This observation suggests that summarizing sensor measurements into hemispheric statistics may reduce the availability of fine-grained spatial information contained within individual MEG channels. In contrast, channel-level representations preserve localized neural activity patterns and sensor-specific characteristics associated with Parkinson’s disease, thereby providing greater discriminative power for classification. These findings support the adoption of channel-level MEG features in the proposed MIRA-Net framework.

### Ablation Study of Fusion Models

To provide a comprehensive evaluation, the Early Fusion, Vanilla DANN, and Baseline SCL frameworks were assessed using Accuracy, Precision, Recall, and F1-score across the Rest, Go, and Passive task conditions. The detailed performance results are presented in the following tables.

**Table 8:** Classification performance of Early Fusion across the a) Rest, b) Go, and c) Passive task conditions.

| Model | Rest |  |  |  | Go |  |  |  | Passive |  |  |  |
| --- | --- | --- | --- | --- | --- | --- | --- | --- | --- | --- | --- | --- |
|  | Accuracy | Precision | Recall | F1-Score | Accuracy | Precision | Recall | F1-Score | Accuracy | Precision | Recall | F1-Score |
| Logistic Regression | 0.7331 | 0.9599 | 0.6445 | 0.7711 | 0.7662 | 0.9717 | 0.6850 | 0.8035 | 0.7424 | 0.9689 | 0.6518 | 0.7792 |
| Random Forest | 0.7776 | 0.8360 | 0.8476 | 0.8417 | 0.8122 | 0.8745 | 0.8535 | 0.8638 | 0.7775 | 0.8401 | 0.8412 | 0.8406 |
| Decision Tree | 0.7482 | 0.8205 | 0.8180 | 0.8192 | 0.7784 | 0.8419 | 0.8404 | 0.8411 | 0.7484 | 0.8206 | 0.8182 | 0.8193 |
| Support Vector Machine (SVM) | 0.7273 | 0.9690 | 0.6294 | 0.7630 | 0.7620 | 0.9784 | 0.6740 | 0.7981 | 0.7339 | 0.9761 | 0.6342 | 0.7687 |
| HistGradientBoosting | 0.7762 | 0.8363 | 0.8447 | 0.8404 | 0.8063 | 0.8877 | 0.8272 | 0.8563 | 0.7780 | 0.8578 | 0.8174 | 0.8371 |
| XGBoost Classifier | 0.7428 | 0.9493 | 0.6671 | 0.7835 | 0.7882 | 0.9664 | 0.7217 | 0.8262 | 0.7506 | 0.9525 | 0.6764 | 0.7909 |
| LightGBM | 0.7511 | 0.9282 | 0.6967 | 0.7958 | 0.7978 | 0.9554 | 0.7444 | 0.8368 | 0.7563 | 0.9364 | 0.6978 | 0.7995 |
| CatBoost | 0.7522 | 0.9273 | 0.6992 | 0.7971 | 0.7967 | 0.9530 | 0.7450 | 0.8362 | 0.7553 | 0.9409 | 0.6922 | 0.7975 |
|  | Accuracy | Precision | Recall | F1-Score | Accuracy | Precision | Recall | F1-Score | Accuracy | Precision | Recall | F1-Score |
| Extra Trees | 0.7664 | 0.8314 | 0.8338 | 0.8325 | 0.8062 | 0.8686 | 0.8502 | 0.8593 | 0.7677 | 0.8355 | 0.8298 | 0.8326 |
| Gradient Boosting | 0.7745 | 0.8345 | 0.8438 | 0.8390 | 0.7981 | 0.8858 | 0.8153 | 0.8489 | 0.7746 | 0.8535 | 0.8167 | 0.8346 |
| Naïve Bayes | 0.7307 | 0.9406 | 0.6548 | 0.7721 | 0.7511 | 0.9822 | 0.6545 | 0.7855 | 0.7208 | 0.9879 | 0.6067 | 0.7516 |
| MLP Classifier | 0.7686 | 0.8362 | 0.8311 | 0.8333 | 0.8056 | 0.8735 | 0.8436 | 0.8579 | 0.7750 | 0.8500 | 0.8234 | 0.8359 |
| Bagging | 0.7699 | 0.8364 | 0.8324 | 0.8344 | 0.8045 | 0.8740 | 0.8405 | 0.8569 | 0.7727 | 0.8420 | 0.8294 | 0.8356 |
| AdaBoost | 0.7549 | 0.7549 | 0.8238 | 0.8249 | 0.7772 | 0.8553 | 0.8199 | 0.8367 | 0.7585 | 0.8663 | 0.7738 | 0.8169 |
| Gradient Boosting (Tuned) | 0.7786 | 0.8340 | 0.8524 | 0.8430 | 0.8057 | 0.8873 | 0.8265 | 0.8558 | 0.7795 | 0.8532 | 0.8262 | 0.8394 |
| LSTM | 0.7755 | 0.8305 | 0.8521 | 0.8409 | 0.7921 | 0.8788 | 0.8143 | 0.8451 | 0.7759 | 0.8635 | 0.8067 | 0.8336 |
| Voting Ensemble<br>(xgb+lgbm+gboost → LR) | 0.7652 | 0.9021 | 0.7438 | 0.8152 | 0.8037 | 0.9413 | 0.7660 | 0.8446 | 0.7661 | 0.9114 | 0.7357 | 0.8141 |
| Stacking Ensemble<br>(xgb+lgbm+cat → LR) | 0.7758 | 0.8423 | 0.8356 | 0.8387 | 0.8096 | 0.8988 | 0.8198 | 0.8573 | 0.7807 | 0.8554 | 0.8253 | 0.8400 |
| Stacking Ensemble<br>(lgbm+cat+mlp → LR) | 0.7524 | 0.9286 | 0.6990 | 0.7975 | 0.8020 | 0.9479 | 0.7581 | 0.8424 | 0.7619 | 0.9367 | 0.7068 | 0.8055 |
| Voting Ensemble<br>(xgb+rf+hgb → LR) | 0.7717 | 0.8622 | 0.8011 | 0.8304 | 0.8074 | 0.9045 | 0.8099 | 0.8544 | 0.7759 | 0.8751 | 0.7922 | 0.8314 |
| Stacking Ensemble<br>(xgb+lgbm+cat → LR) | 0.7758 | 0.8423 | 0.8356 | 0.8387 | 0.8096 | 0.8988 | 0.8198 | 0.8573 | 0.7807 | 0.8554 | 0.8253 | 0.8400 |
| Voting Ensemble<br>(xgb+lgbm+cat → RF) | 0.7551 | 0.9236 | 0.7077 | 0.8013 | 0.7999 | 0.9519 | 0.7514 | 0.8398 | 0.7624 | 0.9346 | 0.7094 | 0.8064 |
| Stacked Ensemble (Bagging<br>+ AdaBoost) | 0.7753 | 0.8373 | 0.8420 | 0.8395 | 0.8085 | 0.8821 | 0.8377 | 0.8592 | 0.7823 | 0.8515 | 0.8336 | 0.8424 |

**Table 9:**
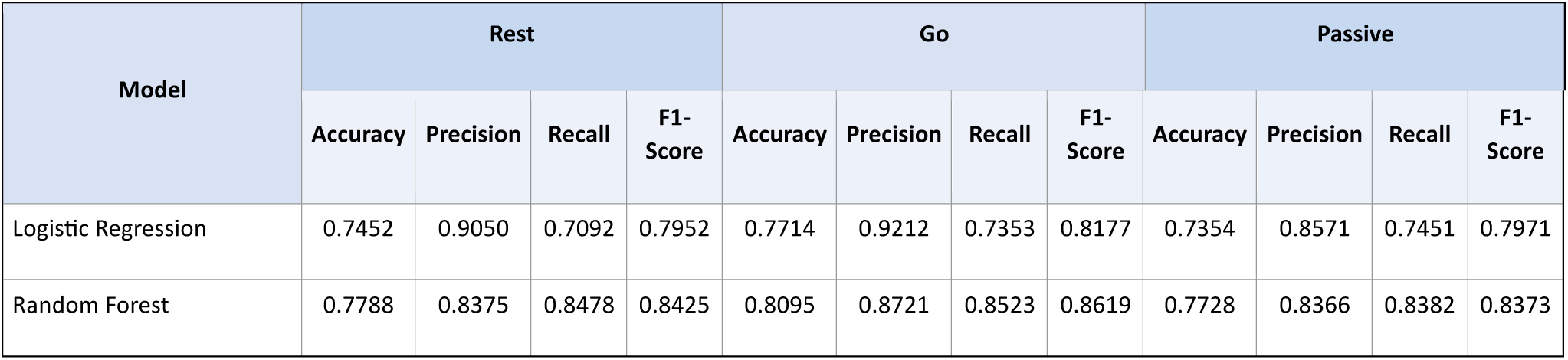
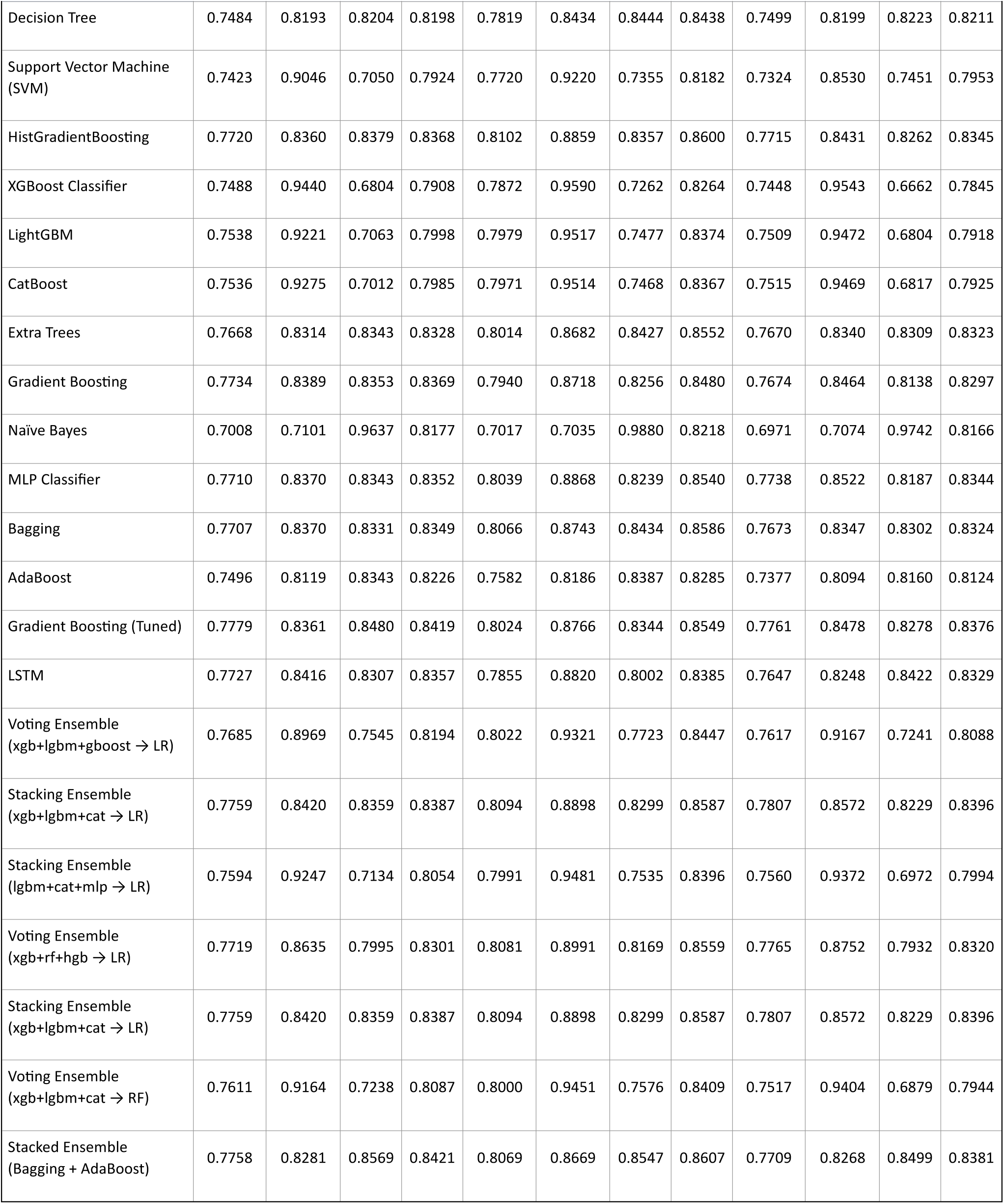
Classification performance of Vanilla DANN across the a) Rest, b) Go, and c) Passive task conditions.

**Table 10:**
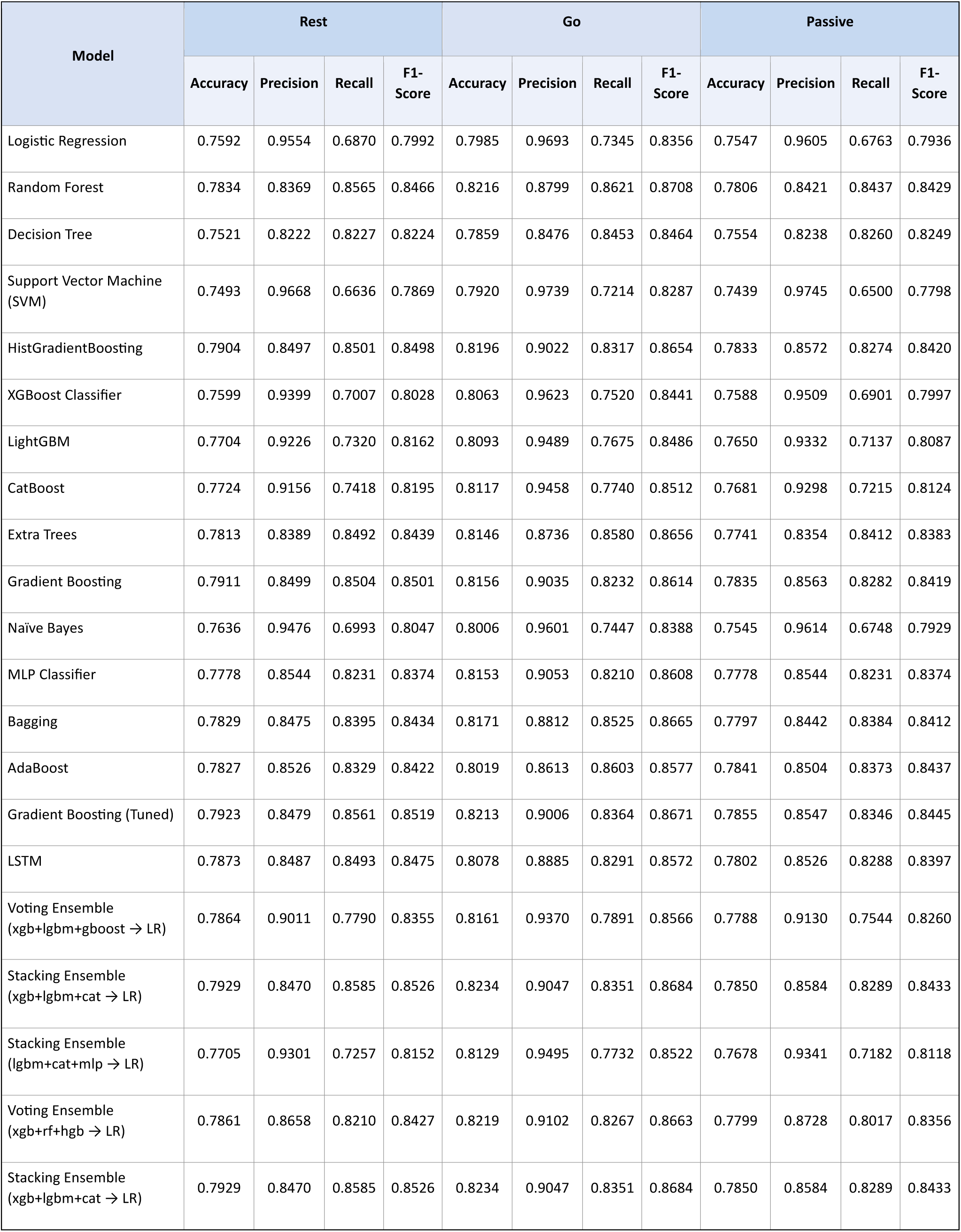
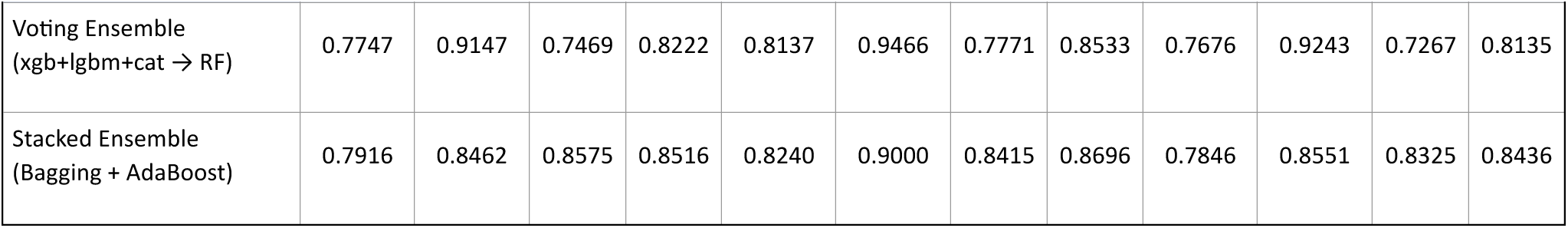
Classification performance of Baseline SCL across the a) Rest, b) Go, and c) Passive task conditions.

| Model | Rest |  |  |  | Go |  |  |  | Passive |  |  |  |
| --- | --- | --- | --- | --- | --- | --- | --- | --- | --- | --- | --- | --- |
|  | Accuracy | Precision | Recall | F1-Score | Accuracy | Precision | Recall | F1-Score | Accuracy | Precision | Recall | F1-Score |
| Logistic Regression | 0.7592 | 0.9554 | 0.6870 | 0.7992 | 0.7985 | 0.9693 | 0.7345 | 0.8356 | 0.7547 | 0.9605 | 0.6763 | 0.7936 |
| Random Forest | 0.7834 | 0.8369 | 0.8565 | 0.8466 | 0.8216 | 0.8799 | 0.8621 | 0.8708 | 0.7806 | 0.8421 | 0.8437 | 0.8429 |
| Decision Tree | 0.7521 | 0.8222 | 0.8227 | 0.8224 | 0.7859 | 0.8476 | 0.8453 | 0.8464 | 0.7554 | 0.8238 | 0.8260 | 0.8249 |
| Support Vector Machine (SVM) | 0.7493 | 0.9668 | 0.6636 | 0.7869 | 0.7920 | 0.9739 | 0.7214 | 0.8287 | 0.7439 | 0.9745 | 0.6500 | 0.7798 |
| HistGradientBoosting | 0.7904 | 0.8497 | 0.8501 | 0.8498 | 0.8196 | 0.9022 | 0.8317 | 0.8654 | 0.7833 | 0.8572 | 0.8274 | 0.8420 |
| XGBoost Classifier | 0.7599 | 0.9399 | 0.7007 | 0.8028 | 0.8063 | 0.9623 | 0.7520 | 0.8441 | 0.7588 | 0.9509 | 0.6901 | 0.7997 |
| LightGBM | 0.7704 | 0.9226 | 0.7320 | 0.8162 | 0.8093 | 0.9489 | 0.7675 | 0.8486 | 0.7650 | 0.9332 | 0.7137 | 0.8087 |
| CatBoost | 0.7724 | 0.9156 | 0.7418 | 0.8195 | 0.8117 | 0.9458 | 0.7740 | 0.8512 | 0.7681 | 0.9298 | 0.7215 | 0.8124 |
| Extra Trees | 0.7813 | 0.8389 | 0.8492 | 0.8439 | 0.8146 | 0.8736 | 0.8580 | 0.8656 | 0.7741 | 0.8354 | 0.8412 | 0.8383 |
| Gradient Boosting | 0.7911 | 0.8499 | 0.8504 | 0.8501 | 0.8156 | 0.9035 | 0.8232 | 0.8614 | 0.7835 | 0.8563 | 0.8282 | 0.8419 |
| Naïve Bayes | 0.7636 | 0.9476 | 0.6993 | 0.8047 | 0.8006 | 0.9601 | 0.7447 | 0.8388 | 0.7545 | 0.9614 | 0.6748 | 0.7929 |
| MLP Classifier | 0.7778 | 0.8544 | 0.8231 | 0.8374 | 0.8153 | 0.9053 | 0.8210 | 0.8608 | 0.7778 | 0.8544 | 0.8231 | 0.8374 |
| Bagging | 0.7829 | 0.8475 | 0.8395 | 0.8434 | 0.8171 | 0.8812 | 0.8525 | 0.8665 | 0.7797 | 0.8442 | 0.8384 | 0.8412 |
| AdaBoost | 0.7827 | 0.8526 | 0.8329 | 0.8422 | 0.8019 | 0.8613 | 0.8603 | 0.8577 | 0.7841 | 0.8504 | 0.8373 | 0.8437 |
| Gradient Boosting (Tuned) | 0.7923 | 0.8479 | 0.8561 | 0.8519 | 0.8213 | 0.9006 | 0.8364 | 0.8671 | 0.7855 | 0.8547 | 0.8346 | 0.8445 |
| LSTM | 0.7873 | 0.8487 | 0.8493 | 0.8475 | 0.8078 | 0.8885 | 0.8291 | 0.8572 | 0.7802 | 0.8526 | 0.8288 | 0.8397 |
| Voting Ensemble (xgb+lgbm+gboost → LR) | 0.7864 | 0.9011 | 0.7790 | 0.8355 | 0.8161 | 0.9370 | 0.7891 | 0.8566 | 0.7788 | 0.9130 | 0.7544 | 0.8260 |
| Stacking Ensemble (xgb+lgbm+cat → LR) | 0.7929 | 0.8470 | 0.8585 | 0.8526 | 0.8234 | 0.9047 | 0.8351 | 0.8684 | 0.7850 | 0.8584 | 0.8289 | 0.8433 |
| Stacking Ensemble (lgbm+cat+mlp → LR) | 0.7705 | 0.9301 | 0.7257 | 0.8152 | 0.8129 | 0.9495 | 0.7732 | 0.8522 | 0.7678 | 0.9341 | 0.7182 | 0.8118 |
| Voting Ensemble (xgb+rf+hgb → LR) | 0.7861 | 0.8658 | 0.8210 | 0.8427 | 0.8219 | 0.9102 | 0.8267 | 0.8663 | 0.7799 | 0.8728 | 0.8017 | 0.8356 |
| Stacking Ensemble (xgb+lgbm+cat → LR) | 0.7929 | 0.8470 | 0.8585 | 0.8526 | 0.8234 | 0.9047 | 0.8351 | 0.8684 | 0.7850 | 0.8584 | 0.8289 | 0.8433 |
|  | Accuracy | Precision | Recall | F1-Score | Accuracy | Precision | Recall | F1-Score | Accuracy | Precision | Recall | F1-Score |
| Voting Ensemble<br>(xgb+lgbm+cat → RF) | 0.7747 | 0.9147 | 0.7469 | 0.8222 | 0.8137 | 0.9466 | 0.7771 | 0.8533 | 0.7676 | 0.9243 | 0.7267 | 0.8135 |
| Stacked Ensemble<br>(Bagging + AdaBoost) | 0.7916 | 0.8462 | 0.8575 | 0.8516 | 0.8240 | 0.9000 | 0.8415 | 0.8696 | 0.7846 | 0.8551 | 0.8325 | 0.8436 |

**Table 11:**
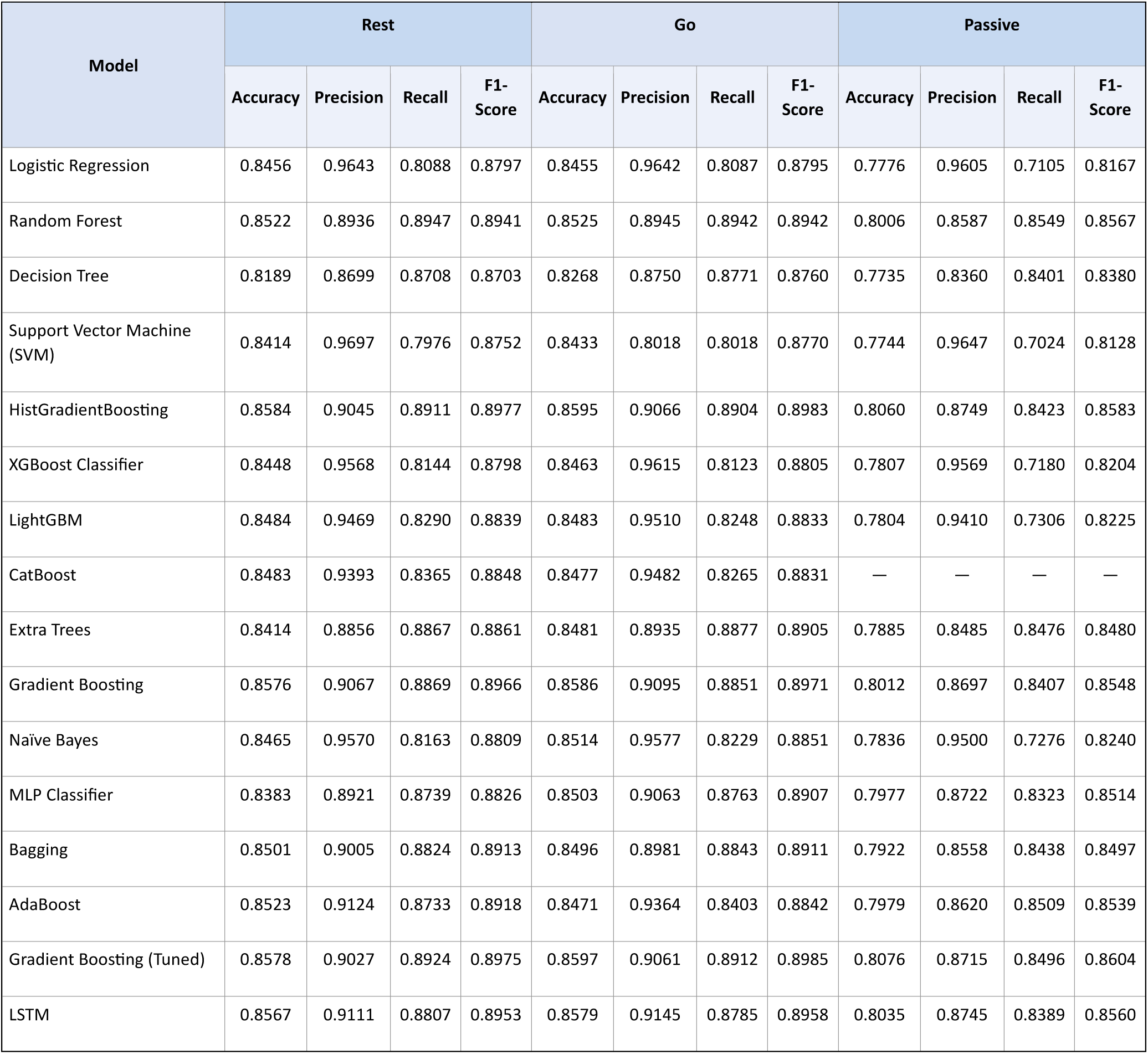
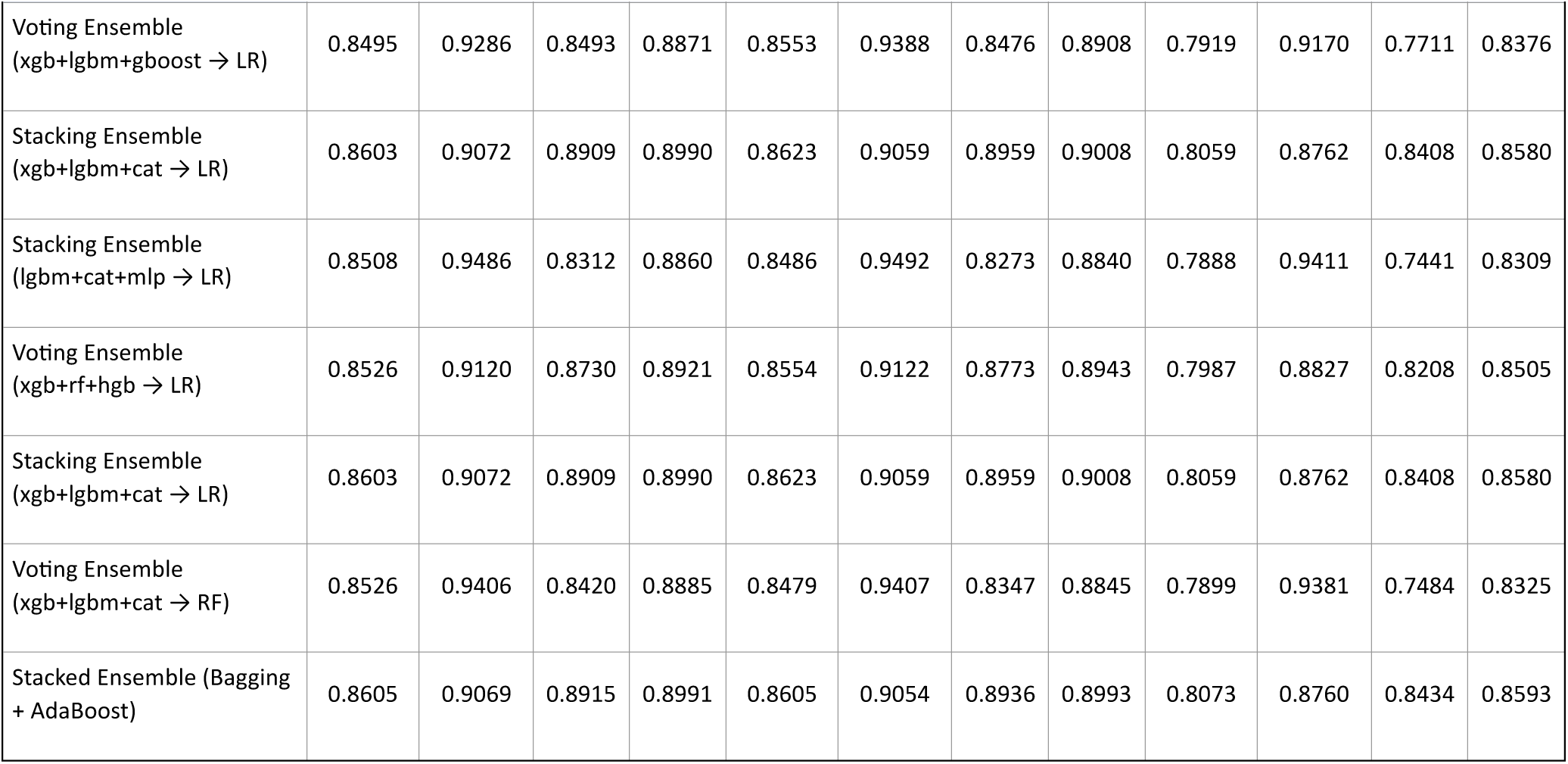
Classification performance of MIRA-Net across the a) Rest, b) Go, and c) Passive task conditions.

From **Tables 8–11**, it is evident that MIRA-Net consistently outperformed Early Fusion, Vanilla DANN, and Supervised Contrastive Learning across the Rest, Go, and Passive task conditions, as evaluated using multiple classifiers and performance metrics, including Accuracy, Precision, Recall, and F1-score. The performance gains were most pronounced under the Go condition while remaining consistently strong under the more challenging Passive condition. These findings further demonstrate the effectiveness of the proposed cross-cohort representation learning strategy in learning robust and discriminative representations for Parkinson’s disease classification.

